# Assessing inhibitory control deficits in adult ADHD: A systematic review and meta-analysis of the stop-signal task

**DOI:** 10.1101/2022.07.09.22277429

**Authors:** Daniel Senkowski, Theresa Ziegler, Mervyn Singh, Andreas Heinz, Jason He, Tim Silk, Robert C. Lorenz

## Abstract

**Background:** In recent years, there has been an increasing quest in improving our understanding of neurocognitive deficits underlying adult attention-deficit/hyperactivity disorder (ADHD). Current statistical manuals of psychiatric disorders emphasize inattention and hyperactivity-impulsivity symptoms, but empirical studies have also shown consistent alterations in inhibitory control. Thus far, there is no established neuropsychological test to assess inhibitory control deficits in adult ADHD. A common paradigm for assessing response inhibition is the stop-signal task (SST).

**Methods:** Following PRISMA-selection criteria, our systematic review and meta-analysis integrated the findings of 26 publications with 27 studies examining the SST in adult ADHD.

**Results:** The meta-analysis, which included 883 patients with adult ADHD and 916 control participants, revealed reliable inhibitory control deficits, as expressed in prolonged SST response times, with a moderate effect size *g* = 0.51. The deficits were not moderated by study quality, sample characteristics or clinical parameters, suggesting that they may be a phenotype in this disorder. The analyses of secondary outcome measures revealed greater SST omission errors and reduced go accuracy in patients, indicative of altered sustained attention. However, only few (N<10) studies were available for these measures.

**Discussion:** Our meta-analysis suggests that the SST could, in conjunction with other tests and questionnaires, become a valuable tool for the assessment of inhibitory control deficits in adult ADHD.

## Introduction

Adult attention deficit hyperactivity disorder (ADHD) is a neurodevelopmental condition that emerges during childhood or young adulthood and is characterized by symptoms of inattention and/or hyperactivity-impulsivity (Adler et al., 2017; Asherson et al., 2016). Adults with ADHD show a global prevalence of 2.58 % (persistent disorder) and 6.76 % (symptomatic disorder) (Song et al., 2021). In clinical practice, adult ADHD is assessed through questionnaires, interviews of relatives and inspection of school certificates. While neurocognitive deficits are inherent in ADHD, there is still no established test or test battery that is generally used in the assessment of this disorder (Fried et al., 2021; Nikolas et al., 2019). Nevertheless, in patients with presumed cognitive deficits, neuropsychological tests should be used to objectify these deficits during the diagnostic process. To date, there is an emerging quest in establishing neurocognitive paradigms as complementary tools in the assessment of adult ADHD.

Early research on ADHD has suggested that deficits in inhibitory control are a primary phenotype in this disorder (Barkley, 1997). Support for the inhibition deficit model comes, among others, from studies that showed altered executive functions in adult ADHD (Hadas et al., 2021; Linhartová et al., 2021; Nigg et al., 2002; Silverstein et al., 2020). An important aspect of executive functions is inhibitory control, which has frequently been investigated with the stop-signal task (SST; Verbruggen and Logan, 2008). In the SST, participants are instructed to perform a forced-choice response following a ‘go-signal’, e.g., an arrow pointing to the left or right, and to respond with a left or right button press, respectively. Crucially, in a small proportion of trials, an auditory or visual ‘stop-signal’ is presented after the go-signal and participants are required to withhold the behavioral response. In most studies an adaptive approach is applied to obtain the go-signal to stop-signal delay interval for which the response inhibition rates are around 50% at an individual subject level. Based on this delay interval and the response times to go-trials, the stop-signal reaction time (SSRT) is calculated, which has become an established measure of response inhibition (Logan et al., 2014). A previous meta-analysis of the SSRT, which involved studies in children and adults diagnosed with ADHD, has revealed deficits of moderate effect sizes across age groups (Lipszyc and Schachar, 2010). This meta-analysis included 60 studies with children but only 10 studies with adult ADHD. Hence, the validity of this analysis regarding adult ADHD was limited and the degree of SSRT deficits in adult ADHD remains to be investigated.

Here, we performed a review and meta-analysis that conformed to current PRISMA-guidelines focusing on response inhibition deficits, as expressed in the SSRT, in adult ADHD. Our analysis included 26 publications with 27 studies, which allowed for a reliable estimation of response inhibition deficits in patients. We conducted a quality assessment of the SST following a recent consensus paper (Verbruggen et al., 2019) and estimated the risk of bias (RoB) for each study. We thoroughly examined if the study quality as well as participant-related and clinical factors influence response inhibition deficits in adult ADHD.

## Methods

The protocol of this systematic review and meta-analysis was pre-registered in the PROSPERO database (PROSPERO ID: CRD42021266709).

### Study selection

The following study selection criteria were applied: 1. Patient population: Included were studies containing at least one group of adult participants (18+) with a current diagnosis of ADHD in accordance with the DSM criteria (5 or earlier) or Hyperkinetic Disorder in accordance with the ICD (10 or earlier) criteria. Studies investigating populations with only subclinical ADHD symptomatology were not considered. 2. Control group: Studies must contain at least one healthy control group. 3. Experimental task: Response inhibition performance had to be obtained by the SST or the Change Task, which is a modified version of the SST in which individuals shift to a secondary response after they have inhibited an ongoing response (Verbruggen and Logan, 2009). Studies using atypical SST paradigms such as dual tasks or the selective SST were excluded. The same applies to studies in which participants received feedback on stop-signal performance, because feedback and reward influence response inhibition (Lipszyc and Schachar, 2010; Slusarek et al., 2001). 4. Outcome measure: Sufficient test statistics for the stop-signal reaction time (SSRT) must be provided to calculate standardized mean differences (Hedge’s g). 5. Other criteria: Empirical articles written in English or German language and published or accepted for publication in peer-reviewed journals during the time range 2000-2022.

### Search strategy

To identify relevant articles, an electronic search was conducted up to April 14, 2022 in two major publication databases: Medline and PsycInfo (accessed from EBSCOhost). The following syntax, adapted from Lipszyc and Schachter (2010), was used: [(attention deficit hyperactivity disorder OR ADHD) AND Adult* AND (stop task OR stop signal OR response inhibition OR executive function)]. Limiters were set to only show articles published in peer-reviewed journals since the 1st of January 2000, in English or German. Furthermore, reference lists of the identified empirical articles, previous meta-analysis and systematic reviews were scanned to ensure that all relevant articles were captured.

### Study selection

The study selection process was conducted by two authors (TZ and DS) and included two stages: 1. Initial screening of titles and abstracts using the above-described inclusion and exclusion criteria. 2. For the resulting set of studies full texts were obtained and checked in detail for eligibility. Screening of eligible articles was performed in Endnote. In case of discrepancies between authors regarding the eligibility of studies, studies were screened by other team members and disagreements during the first or the second screening process were discussed until consensus was reached.

### Data extraction and outcomes

Data was extracted by two authors independently (TZ and MS). If statistical values for the meta-analysis were not sufficiently reported, the authors of the articles were contacted and asked to provide the missing information. The following measures of the SST were extracted from the articles, separately for patients and controls: SSRT as primary outcome; stop commission errors (responding on a stop trial); go discrimination errors (e.g., responding with the left arrow key, even though a rightwards pointing arrow was presented); go omission errors (not responding on a go trial) and go accuracy (percentage of correct go trials) as secondary outcomes. In addition, important variables that could influence behavioral performance in the planned analysis were extracted and tabulated for each study: Age; IQ; percentage of males; ADHD subtype; years of education; comorbidities; medication status; recruitment setting of patients.

### Assessments of the SST validity and risk of bias

The validity of the SST and the risk of bias (RoB) in each study are important factors that could influence group differences between patients and controls. Therefore, they were explicitly examined in the current analysis. Both SST validity and RoB were assessed by two independent raters (TZ and MS). In case of discrepancies between authors, consensus was reached from discussions with other team members. To increase inter-rater reliability, a calibration session was conducted in which the assessments were applied to two articles not included in this review. This way, the two raters could identify possible sources of disagreement and decide on rules on how to rate ambiguous cases (*Supplementary text 1*).

Across studies selected for this meta-analysis, there is considerable variability in the administration of the SST, and the analytical procedures used to derive outcome measures. To determine SST validity, we used the recent consensus guide developed by Verbruggen et al. (2019), which offers 12 ‘best practice’ recommendations on how to design, implement and analyze the SST. We selected four main criteria from this consensus guide and rephrased them into four dichotomous items, i.e., item fulfilled or not, for the critical appraisal (*Supplementary text 2*). In case of missing information, the criterion was rated as not fulfilled.

The overall SST validity was then rated as follows: <3 criteria fulfilled = low validity; 3 criteria fulfilled = moderate validity; 4 criteria fulfilled = high validity. Cohen’s unweighted kappa for nominal data were calculated for each item. In case of a bias or a prevalence problem, Byrt’s bias and prevalence adjusted kappa were additionally reported (Byrt et al., 1993; Hallgren, 2012). For the RoB assessment, we applied the adapted Hombrados and Waddington criteria that have been recently used for studies with ADHD patients (Hulsbosch et al., 2021): (1) Equivalent group sizes; (2) Use of a diagnostic interview or questionnaires to determine ADHD diagnosis; (3) Sufficient sample sizes; (4) All statistical outcomes are reported; (5) Transparent report of the data analysis; (6) Report of missing/excluded data. Each item was rated “good/low RoB”, “satisfactory/moderate RoB”, or “poor/high RoB”. In accordance with the rating system described in the Cochrane Handbook, an overall quality rating of low, moderate or high was applied to each study based on the following criteria: If at least one of the categories was rated as having a moderate RoB, the overall RoB could only be rated as moderate as well, even if all other categories were considered having a low RoB. The same principle applied when at least one category was appraised as having a high RoB. Cohen’s weighted Kappa was calculated for each individual domain (Cohen, 1968; Hallgren, 2012). After all studies were rated regarding SST validity and RoB, a variable of overall study quality combining the RoB ratings and SST validity was created. Studies with high RoB and low SST validity were rated as having low overall quality, studies with moderate or low RoB AND moderate or high SST validity were rated as having moderate to high overall quality. Studies characterized with the remaining combinations of RoB and SST validity (low RoB and low SST validity; moderate RoB and low SST validity; high RoB and moderate SST validity; high RoB and high SST validity) were designated to the category moderate to low overall quality. This categorization was used for subgroup analysis (see below).

### Meta-analysis

The meta-analysis was carried out in R (version 4.0.3; R Core Team, 2020) and the metafor package (version 3.0.2; Viechtbauer, 2010). Hedges’ *g* was calculated for each individual study and each outcome (primary outcome SSRT and secondary outcomes) displaying the effect size of the group difference. Given that various sources could account for differences in findings across studies, e.g., examination of different patient samples or use of different SST paradigms, a random-effects model was fitted to the data. Instead of the usual large-sample approximation, the sampling variance was adjusted by taking the sample-size weighted average of the Hedges’ *g* values into the equation, as this approach has been shown to be less biased (Lin and Aloe, 2021). For computing confidence intervals, the method introduced by Knapp and Hartung (2003) was chosen. To assess for heterogeneity, (1) τ^2^ was estimated using the restricted maximum-likelihood estimator (Viechtbauer, 2005), (2) the *Q*-test for heterogeneity and (3) the *I*^2^ statistics (Higgins and Thompson, 2002) are reported. If heterogeneity between studies is present, i.e., τ^^2^ > 0, regardless of whether the *Q*-test reaches significance, a prediction interval for the true outcomes is provided (Riley et al., 2011). The results will be visualized using forest plots. Furthermore, the model is assessed regarding (1) potential outliers, i.e. studies with studentized residuals larger than the 100 × (1 − 0.05/(2 × *k*))th percentile of a standard normal distribution, considering a Bonferroni correction with *α* = 0.05 (two-sided) for k included studies as well as (2) potentially overinfluential studies, i.e. with a Cook’s distance larger than the median plus 6 times the interquartile range of the Cook’s distances (Viechtbauer and Cheung, 2010). If outliers were detected, leave-one-out diagnostics for sensitivity analysis were conducted.

### Assessment of publication bias

Evidence of publication bias was assessed using a combination of visual and statistical approaches. First, the funnel plot (Copas and Chi, 2000) of standardized mean difference (SMD) against the inverse square root of the sample size was visually inspected for asymmetries (Zwetsloot et al., 2017). If no bias exists, the funnel plot should be symmetrical and narrow down at the top, where studies with larger sample sizes are located and estimation of the effect is more precise. However, determination of publication bias using visual inspection methods (such as funnel plots) are often subjective and prone to judgmental errors (Wang and Bushman, 1998). Therefore, it is recommended to additionally compute a quantile-quantile plot (Q-Q plot) to aid in the assessment of publication bias. Next, Egger’s regression test was calculated using the inverse of the square root sample size as a predictor to statistically test for asymmetry of the funnel plot (Zwetsloot et al., 2017). Where evidence of bias exists, the trim-and-fill method was applied to adjust for publication bias (Shi and Lin, 2019).

### Meta-regression and subgroup analysis

To assess whether the pre-specified extracted demographic and clinical variables as well as study quality influenced the meta-analytic outcome and to explore the cause of potential heterogeneity, a meta-regression analysis was conducted for continuous (age, sex, IQ) and a subgroup analysis for categorical covariates (RoB, SST validity and overall study quality, comorbidities, patient setting and medication status).

Mixed-effect models were fitted to the data for the meta-regression analysis. If enough data were available across studies, the extracted variables were included in a multivariate regression model. Otherwise, univariate models were estimated. The parameter τ^2^, which displays the residual heterogeneity not explained by the included moderators (Viechtbauer, 2010) was estimated using the REML-estimator (Viechtbauer, 2005). Tests and confidence intervals were calculated by the Knapp and Hartung (2003) method. The mean values for age, IQ, and percentage of males of the study samples were computed to be included in a regression model. To this end, the reported means of patients and the means of controls were averaged. If sample sizes of ADHD participants and healthy controls differed substantially, a sample-size weighted mean was calculated. If IQ values were reported for verbal and non-verbal IQ separately in a study, the average of those values was calculated. IQ was centered before taken into a univariate regression model. Age and gender were standardized before taken into a multivariate regression model.

The following variables were considered for subgroup analysis: (1) RoB with the subgroups low RoB, moderate RoB and high RoB; (2) SST validity with the subgroups low validity, moderate validity and high validity; (3) overall study quality with the subgroups low overall quality, low to moderate overall quality and moderate to high overall quality; (4) comorbidities in patients with the subgroups comorbidities in patients allowed and comorbidities in patients not allowed; (5) comorbidities in controls with the subgroups comorbidities in controls allowed and comorbidities in controls not allowed; (6) patient setting with the subgroups recruited from a clinical-setting, recruited from a non-clinical setting and recruited from both (mixed) and finally (7) medication status with the subgroups medicated and unmedicated. For each of these variables, separate random-effects models were fitted for each subgroup. Then, a fixed-effects regression, including a moderator containing the subgroups’ effect estimates, was calculated to test whether it significantly moderated SSRT.

Finally, for both meta-regressions and subgroup analyses an omnibus test of moderators was conducted, testing all coefficients excluding the intercept against 0. If the omnibus test reaches significance, this might be an indication that some of the heterogeneity could be explained by the predictors included in the model (Viechtbauer, 2010).

### Secondary outcome measures of the SST

In addition to the SSRT, the SST reveals other outcome measures that are recommended to be reported (Verbruggen et al., 2019). For the current meta-analysis, we examined stop commission errors, go discrimination errors, go omission errors and go accuracy. All studies which reported these parameters were used for the meta-analyses. The procedures were the same as for SSRT, except that no meta-regression or subgroup analyses were conducted, due to the lower number of available studies.

## Results

### Study selection

The electronic search resulted in 1186 articles in MEDLINE and 1353 in PsycInfo (*Figure 1*). Limiters described in the methods section excluded 215 of these articles. Search results were exported into EndNote, during which EBSCOhost automatically removed 662 duplicates, resulting in 1662 studies. After removing the remaining duplicates using EndNote’s automatic deduplication tool (n = 177) and manual inspection (n = 36), there were 1449 remaining articles. Screening of titles and abstracts for eligibility led to the exclusion of 1288 studies. For the remaining 161 studies, full texts were obtained and checked thoroughly for eligibility. From the 161 studies, 116 were excluded because they did not include a healthy control group (n = 1), only subclinical symptomatology was assessed (n = 2), ADHD was only included as comorbid disorder (n = 1) or no SST paradigm was used (n = 112). A further 4 studies were excluded because they had substantially modified the SST paradigm and another 8 studies because their samples included both children and adults. In six studies not sufficient statistical values for the meta-analysis were reported and authors were contacted. We received data from four studies, which were then included in the final sample. Finally, it is important to note that Bekker et al. (2005a, 2005b) and van Dongen-Boomsa et al. (2010) reported data from the same experimental session and the same participant sample. The same accounted for Nigg et al. (2005), Stavro et al. (2007) and Martel et al. (2017) as well as for Linhartová et al. (2020) and Linhartová et al. (2021). For these groups of articles, the reported values were extracted and counted as one study in the meta-analysis. Finally, Szekely et al. (2017) conducted two SST experiments, one implemented for fMRI and one for MEG. Even though samples for these two experiments partially overlap (63 completed the SST during MEG and fMRI, 85 during fMRI only, and 33 during MEG only), they were treated as individual observations in the analysis. The screening of reference lists revealed no further articles. Finally, there were a total of 26 publications with 27 studies included in the meta-analysis (1799 participants; ADHD = 883; controls = 916). Sample characteristics for all included studies are found in *Table 1*.

**Figure 1:**
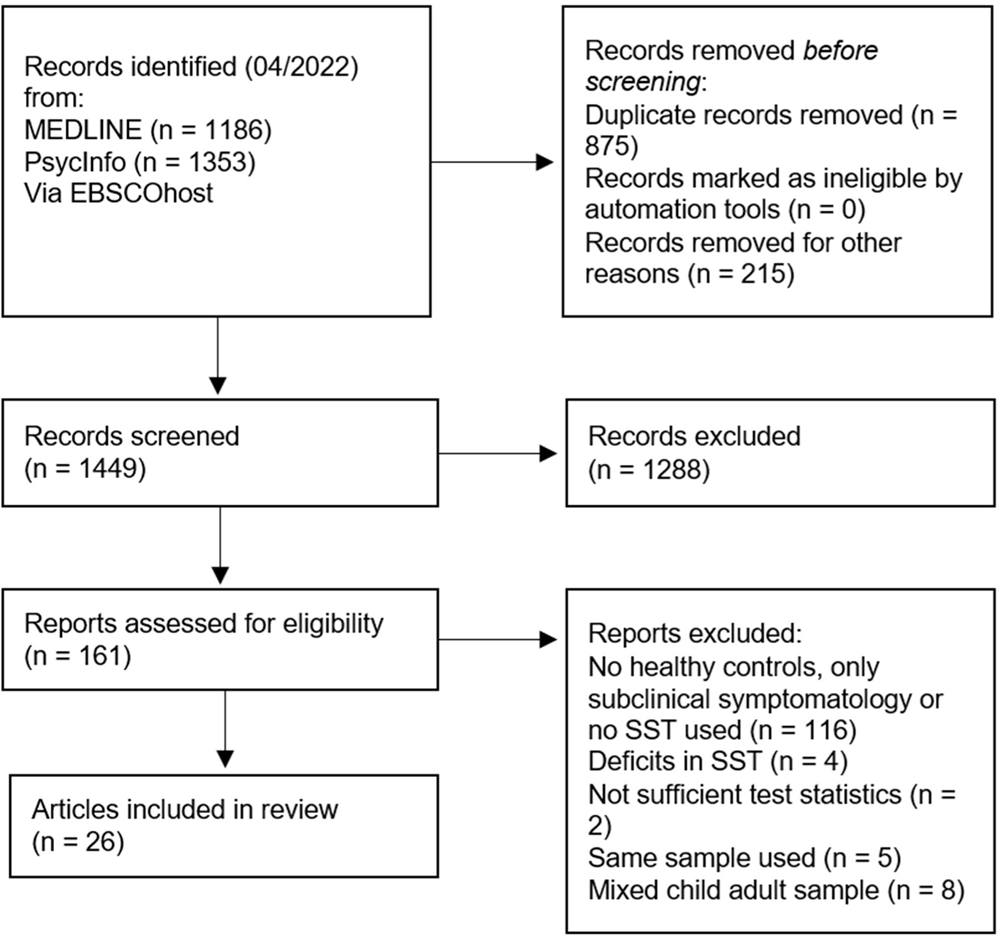
PRISMA chart of study selection (in accordance with Page et al., 2021)

**Table 1:**
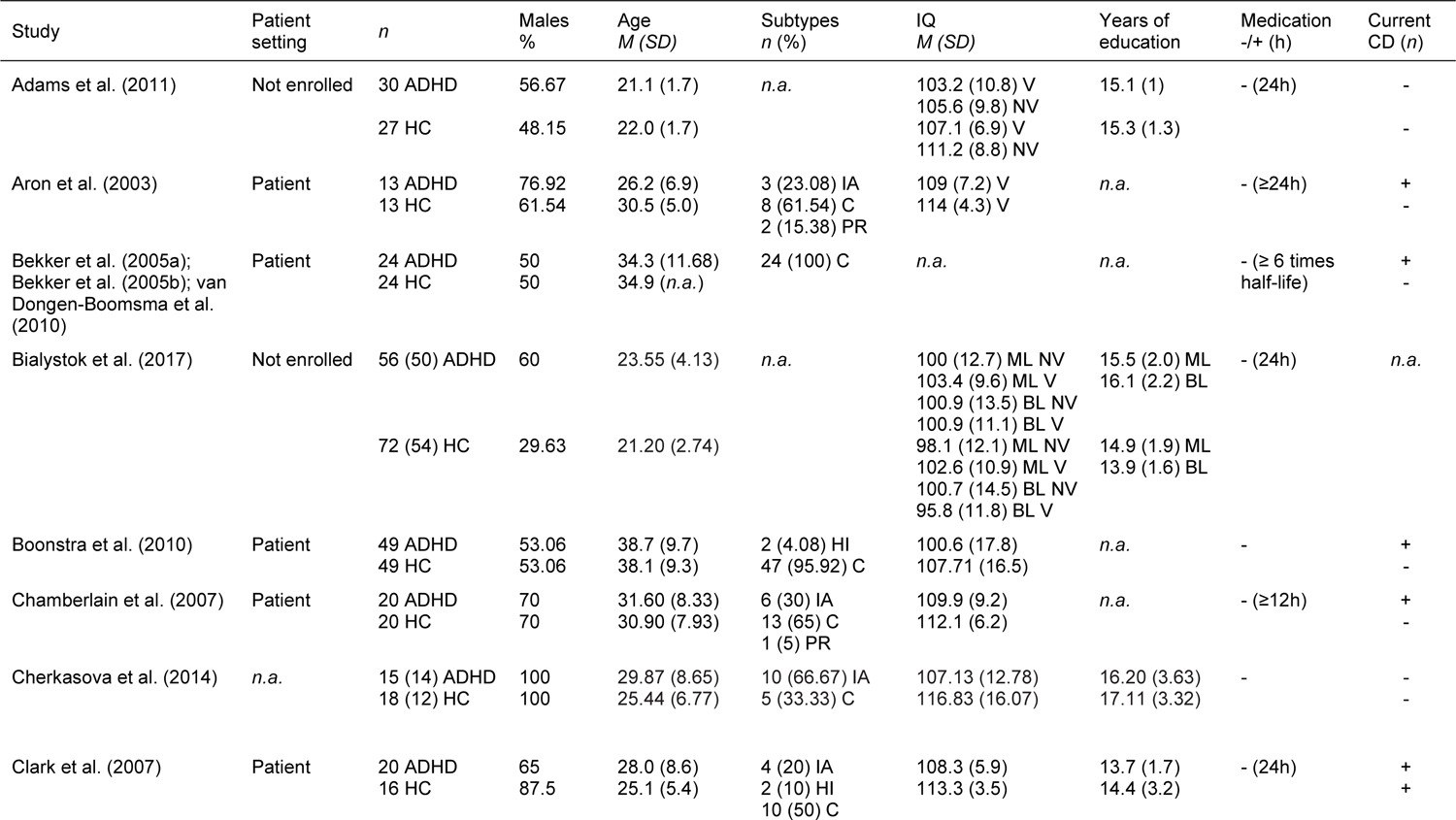
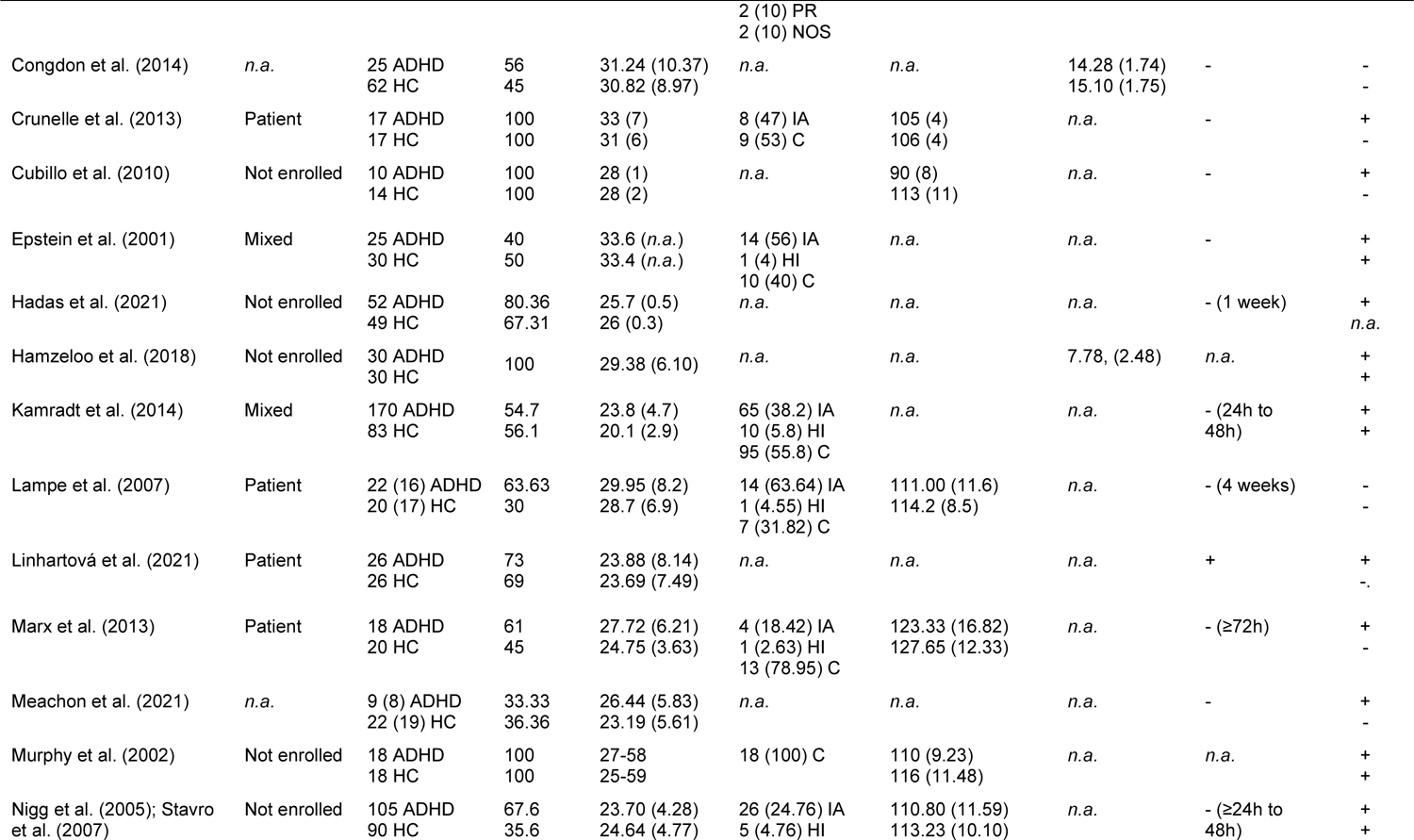
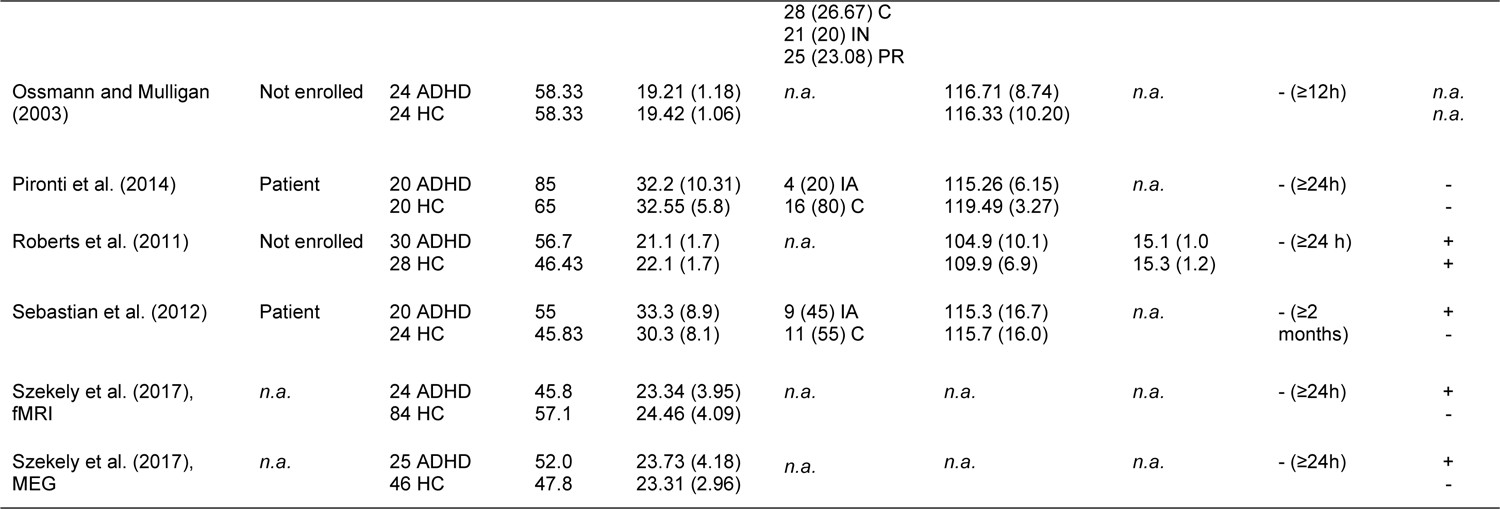
Studies investigating the stop-signal task in adult ADHD. ADHD: attention-deficit hyperactivity disorder; HC: healthy controls; Patient Setting: ADHD group recruited from in/outpatient setting or non-clinical setting; Years of education: If education status was presented in any other form than years of education, it was not included in this table. Medication: allowed or not allowed during testing (+ and -, respectively). If medication was not allowed, duration of omission prior to testing in brackets, if medication was allowed, percentage of medicated participants in brackets; CD: comorbid disorder; V: verbal; NV: nonverbal; ML: monolingual; BL: bilingual; IA: predominantly inattentive subtype; HI: predominantly hyperactive-impulsive subtype; C: combined subtype; PR: in partial remission; NOS: Not otherwise specified; IN: inconsistent, met criteria for ADHD–H, ADHD–C, or ADHD–I as children but for a different subtype as an adult.

Twenty-four out of 27 studies prohibited stimulant medication on the day of testing, two studies did not report this information and one study allowed medication (Linhartová et al., 2021). One study tested the effect of stimulant medication on task performance (Chamberlain et al., 2007) and another study allowed medication during testing (Congdon et al., 2014). To maintain similarity between studies, data of those two articles was extracted for the placebo, i.e., the unmedicated group only. Marx et al. (2013) used an SST paradigm that compared performance with and without reward. For this study only data for the non-reward group were extracted. In some articles, information on the presence of comorbidities or the patient setting was ambiguously reported. For example, Aron et al. (2003) report that healthy controls had “no previous contact with psychiatric services” but it is unclear whether potential comorbidities of healthy controls were screened within the study. Therefore, the coding for those two variables might be biased. Upon request, Meachon et al. (2021) provided non-published information on the age and sex distribution in the two groups. Bialystok et al. (2017) provided the means of age and male percentage for the subset that completed the SST. Demographic variables, information about the IQ and other relevant information about the study population were not available for all studies. A summary is given in *Table 2*.

**Table 2:** Sample characterization. n: number of participants, k: number of studies reporting this information; Males: percentage of males in the sample; clinical: recruited from a clinical (inpatient/ outpatient) setting; non-clinical: recruited from a non-clinical setting (e.g., newspaper, university); mixed: recruited both from a clinical and a non-clinical setting; In partial remission: met at least 6 of 9 DSM-5 criteria in childhood, but only 3 to 5 of 9 criteria in adulthood. Inconsistent: met the diagnostic criteria for a different subtype in childhood than in adulthood; NOS: not otherwise specified.

|  | ADHD patients | Healthy controls | Total |
| --- | --- | --- | --- |
| *N* | 883 | 916 | 1799 |
| Age (*k* = 26) | 27.73 (4.92) | 26.98 (4.92) | 27.44 (4.72) |
| Males, % (*k* = 27) | 67.19 (19.53) | 61.30 (22.66) | 64.25 (20.08) |
| IQ (*k* = 17) | 108.41 (7.48) | 113.17 (6.08) | 110.85 (6.12) |
| Patient Setting, *k* |  |  |  |
| Clinical | 11 | *n.a.* | 11 |
| Non-clinical | 9 |  | 9 |
| Mixed | 2 |  | 2 |
| Comorbidities, *k* |  |  |  |
| Allowed | 20 | 7 | *n.a.* |
| Not allowed | 5 | 17 |  |
| Subtypes, *n* (*k* = 15) |  |  |  |
| Primarily Inattentive | 167 | *n.a.* | 167 |
| Primarily Hyperactive | 314 |  | 314 |
| Combined | 22 |  | 22 |
| In Partial Remission | 30 |  | 30 |
| Inconsistent | 21 |  | 21 |
| NOS | 2 |  | 2 |

### SST validity and risk of bias

SST validity was evaluated for all 26 articles across 4 items, resulting in 104 individual ratings. *Table 3* provides an overview of the ratings. Nineteen studies (73%) were rated low validity, 5 studies (19%) moderate and 2 studies (8%) as high validity. Marginal distributions revealed a prevalence bias for all items to some degree. This bias was strongest for item 2 and item 4. All items showed substantial to perfect interrater agreement (Hallgren, 2012) (*Supplementary Table 1*). Taken together, most studies included in the meta-analysis had a low or moderate quality of the SST.

**Table 3:**
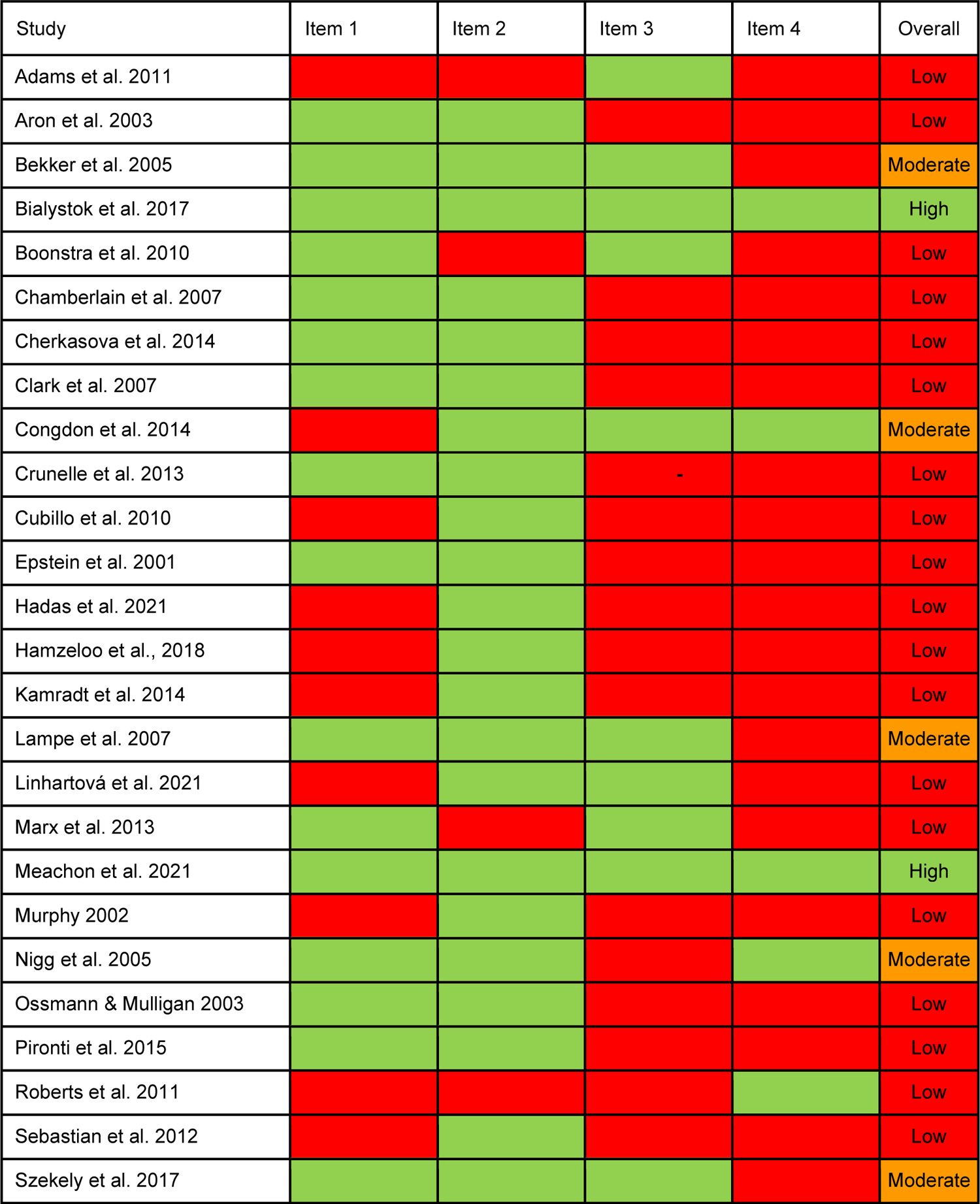
Stop-signal task validity ratings. Item 1: ≥50 stop trials in total, stop trials constituting ≤25% of all trials; Item 2: staircase algorithm implemented; Item 3: integration method used; Item 4: Cut-Offs applied to ensure valid SSRT estimation; green: fulfilled; red: not fulfilled; Low: 1 or 2 out of 4 items fulfilled; Moderate: 3 out of 4 items fulfilled; High: 4 out of 4 items fulfilled.

RoB was evaluated for all 26 articles across 6 domains, resulting in 156 individual ratings. *Table 5* provides an overview of the RoB ratings. One article (4%) received a low rating, Twelve articles (46%) a moderate rating and 13 articles (50%) a high rating. For all domains, there was substantial to perfect interrater agreement (*Supplementary Table 2*). Overall, most articles had a moderate or high RoB.

**Table 4:**
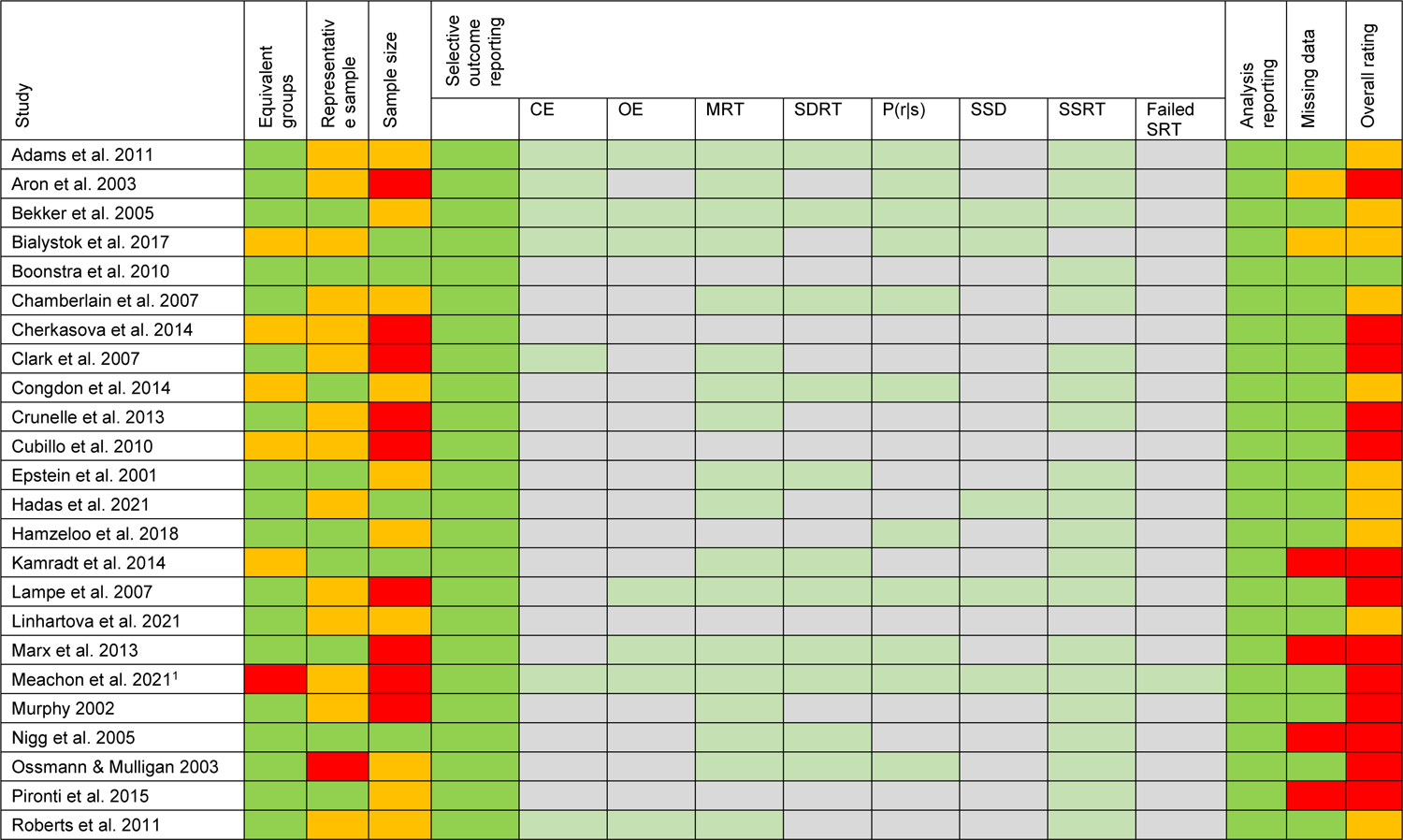
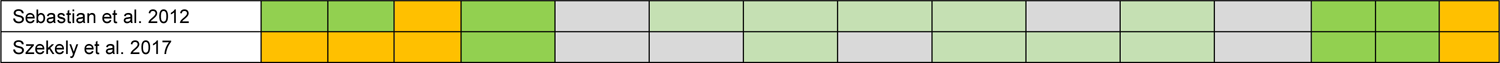
*Risk of bias ratings*. Ratings were based on the adapted Hombrados and Waddington criteria (Hulsbosch et al., 2021); CE: choice errors; OE: omission errors; MRT: mean reaction time; SDRT: intrasubject variability; P(r|s): probability to respond on a stop trial; SSD: stop signal delay; SSRT: stop signal reaction time; Failed SRT: failed stop reaction time; green: good/low RoB; orange: satisfactory/moderate RoB; red: poor/high RoB. ^1^This study was a pilot study, which might be the reason for small sample sizes.

**Table 5:** Meta-regression analyses for SSRT. k: number of studies for which data was available; B: regression coefficient. For categorical variables, B is the average estimated effect size for each individual factor level; SE: standard error of regression coefficient; t: t-test for the regression coefficient; p: p-value for regression coefficient t-test; ci: confidence interval; F-Test: omnibustest of moderator; p_F_: p-value for test of moderator; Sex: percentage of males in the individual study samples; IQ: for ADHD and control group combined; Setting: patient setting of ADHD group.

| Moderator | *B* (*SE*) | *t* | *p* | *ci* | *F*-Test | *p<sub>F</sub>* |
| --- | --- | --- | --- | --- | --- | --- |
| Age, Sex (*k* = 25) |  |  |  |  | *F*(3,21) = 0.558 | .649 |
| Intercept | 0.513 (0.056) | 9.180 | <.001 | 0.397, 0.629 |  |  |
| Age | 0.086 (0.078) | 1.097 | .285 | -0.077, 0.248 |  |  |
| Sex | -0.003 (0.072) | -0.038 | .970 | -0.153, 0.147 |  |  |
| Age:Sex | 0.055 (0.116) | 0.470 | .643 | -0.187, 0.296 |  |  |
| IQ (*k* = 17) |  |  |  |  | *F*(1,15) = 0.384 | .545 |
| Intercept | 0.602 (0.079) | 7.586 | <.001 | 0.433, 0.771 |  |  |
| IQ | 0.008 (0.013) | 0.620 | .545 | -0.020, 0.036 |  |  |

### Meta-analysis of stop-signal reaction time

*Figure 2* presents the forest plot of the observed group differences in the SSRT for 27 observations. Across studies, Hedges’ *g* values ranged from −0.341 to 1.230. Results of the random-effects meta-analysis revealed a statistically significant moderate average effect size estimate of 0.509 (*t*(26) = 7.829, *p* < 0.0001, 95% CI: 0.376-0.644). Adults with ADHD showed moderately higher SSRTs compared to healthy controls. The *I*^2^ statistic demonstrated moderate evidence of heterogeneity across studies (*Q*(26) = 39.546, *p* = 0.043, τ^^2^ = 0.030, *I*^2^ = 31.224%). The heterogeneity reflects in a 95% prediction interval ranging between 0.129 and 0.891.

**Figure 2:**
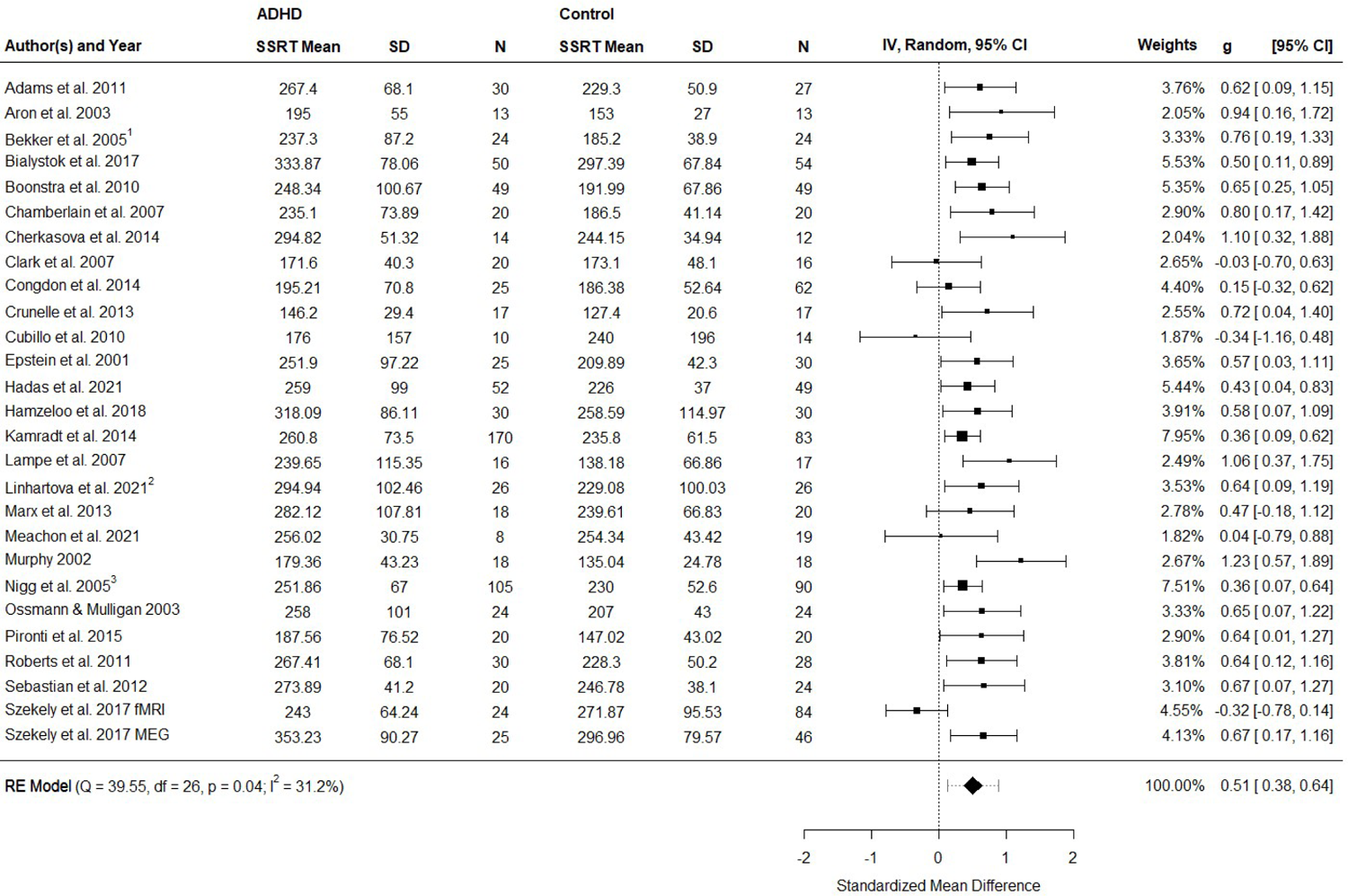
Forest plot showing the observed standardized mean differences (Hedges’ g) for SSRT and the estimate of the random-effects model. The dashed line at the overall effect estimate (diamond) represents the prediction interval that is shown due to the present heterogeneity. ^1^Data was extracted from Bekker et al. (2005a), Bekker et al. (2005b) and van Dongen-Boomsma et al. (2010); ^2^ Data was extracted from Linhartová et al. (2020) and Linhartová et al. (2021); ^3^Data was extracted from Nigg et al. (2005), Stavro et al. (2007) and Martel et al. (2017).

According to the Cook’s distances, none of the studies was overly influential. However, the study of Szekeley et al. (2017) implementing the SST for fMRI had a studentized residual larger than ±3.113 and, hence, is an outlier in the context of this model. Leaving out this observation would reduce τ^^2^ to 0.000, *I*^2^ to 0.004% and increase *g* to 0.524 (95% CI 0.416 to 0.631). Taken together, the random-effects meta-analysis revealed moderate effect sizes (*g* = 0.51 to 0.52) with larger SSRTs in patients compared to controls.

### Publication bias

*Figure 3A* depicts a funnel plot of the studies’ SMDs plotted against the inverse of the square root of the sample sizes. Egger’s regression test for funnel plot asymmetry was not significant (*t*(25) = 1.941, *p* = 0.064). The funnel plot seems to converge close to the average estimate with increasing sample size. A normal quantile-quantile plot is shown in *Figure 3B*. Most dots in this plot fall inside the 95%-confidence bands. However, in the middle of the line there is a slight skewing to the left, with several dots outside the bands. This is an indication that there could be a subtle publication bias. For exploratory purposes, a funnel plot adjusted for publication bias using the trim-and-fill method was computed (*Figure 3C*). The adjusted average effect size estimate is *g* = 0.439, which is still highly significant (*t*(31) = 6.304, *p* < 0.0001, 95% CI: 0.297-0.581). Hence, even if the trim-and-fill method is applied, patients show significantly longer SSRTs compared to controls.

**Figure 3:**
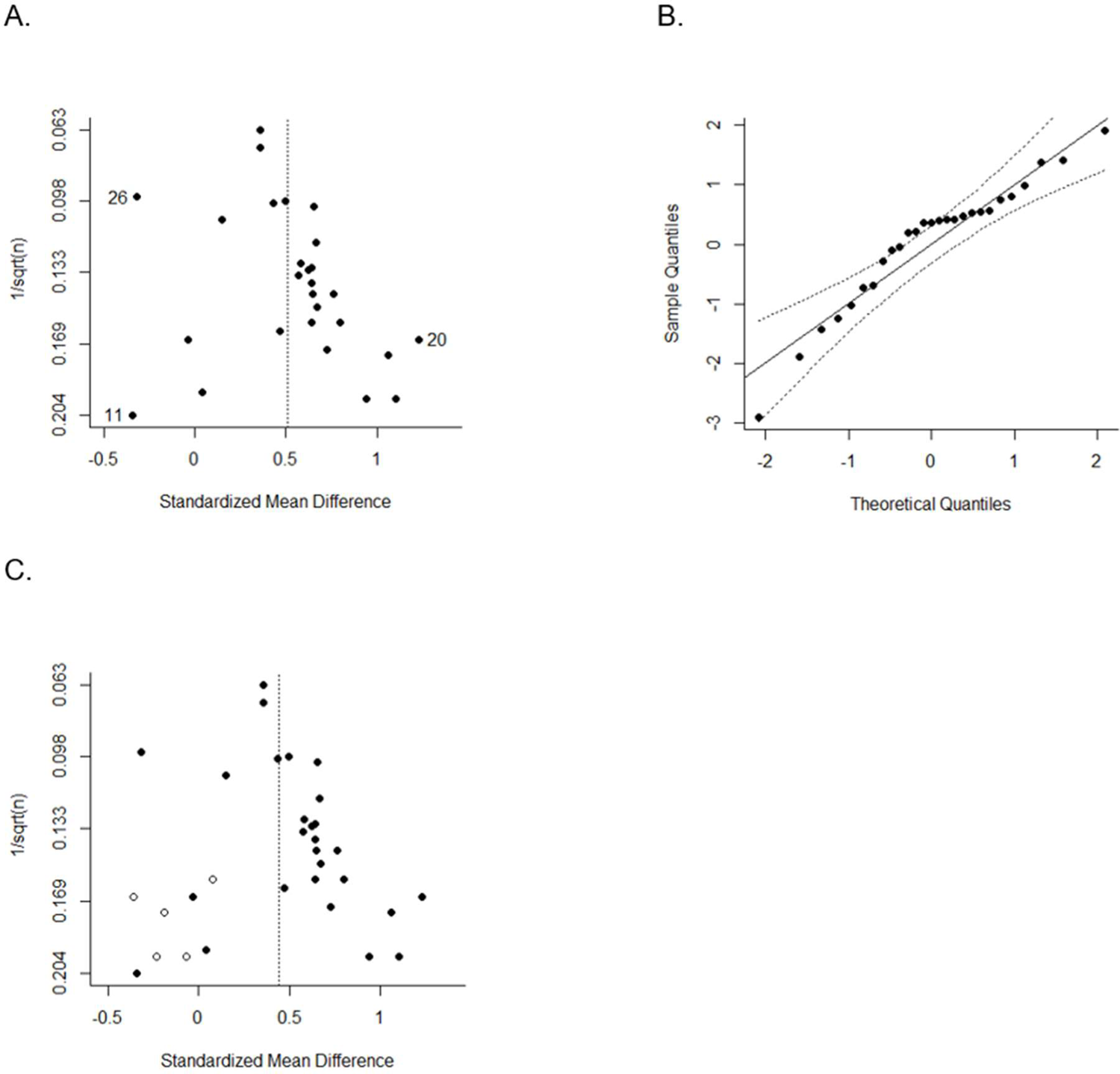
Plots for assessment of publication bias. (A) Funnel plot for SSRT plotting SMDs against the inverse of the square root of the sample size. (B) Normal quantile-quantile plot, plotting the quantiles of a standard normal distribution against the quantiles of the observed distribution. The points should fall on a straight line and inside the 95%-confidence bands. (C) Funnel plot for SSRT plotting SMDs against the inverse of the square root of the sample size adjusted for publication bias using the trim-and-fill method (filled dots: observed effect estimates, non-filled dots: imputed effect estimates). ^11^Cubillo et al. (2010), ^20^Murphy et al. (2002), ^26^Szekely et al. (2017).

### Meta-regression and subgroup analysis

Meta-regression analysis was conducted for continuous covariates (age; sex; IQ) and a subgroup analysis for categorical covariates (RoB, SST validity; overall study quality; comorbidities; patient setting; medication status). In this analysis, the data from the fMRI study by Szekeley et al. (2017) were an outlier and therefore, they were excluded from further analysis. In four of the studies (Bialystok et al., 2017; Cherkasova et al., 2014; Lampe et al., 2007; Meachon et al., 2021) only a subset of participants completed the SST. To assess whether this might influence robustness of meta-regression results, the analysis was conducted again, excluding these 4 studies. This did not substantially influence the study outcome (*Supplementary Tables 3-4*). *Table 5* provides an overview of the meta-regression analyses. Bialystok et al. (2017) reported demographic and outcome variables separately for monolinguals (ML) and bilinguals (BL) in each group. Values for ML and BL were averaged to obtain only a single value per group to be included in meta-regression analysis. The analysis did not reveal any significant effects of age, sex or IQ. *Table 6* provides an overview of the subgroup analyses. Some studies reported that only some psychiatric comorbidities lead to exclusion. Those were coded as “comorbidities allowed” as well. Data for years of education was sparse and heterogeneous and therefore, not included in the meta-regression. As only one study reported that participants were medicated during testing, medication status was also dropped from analysis. There were only 5 articles that did not allow comorbidities in ADHD patients. Therefore, the estimated average SMD for these studies reported in *Table 3* may not be robust. The same accounts for the estimated SMD for the level “mixed” of the setting variable, as only 2 studies report to have recruited ADHD patients from clinical as well as from non-clinical settings. The analysis of study quality revealed that both RoB assessment and SST validity ratings did not significantly moderate SSRT. For RoB, the estimated effect was largest for studies with low RoB (*g* = 0.651) and smallest for studies with high RoB (*g* = 0.531). However, there was only one study designated as having a low RoB, therefore the result for this category should be interpreted with caution. The group of studies assigned low SST validity showed the largest average effect size (*g* = 0.556), whereas the group rated as having high SST validity showed the smallest average effect size (*g* = 0.415). There were only two studies with high SST validity, limiting the reliability of the result for this category. Similar to RoB, the study quality did not significantly moderate SSRT, with an effect size *g* = 0.49 for studies with moderate to high overall quality ratings. Forest plots with subgroups are shown in *Supplementary Figures 1-3*. In summary, our analysis did not reveal variables that significantly moderated the SSRT deficits in adult ADHD.

**Table 6:** Subgroup analysis for SSRT. k: number of studies for which data was available; B: regression coefficients (first group is the intercept, for the other groups the coefficients are contrasts); SE: standard error of regression coefficient; Wald-type z-test for the regression coefficient; p: p-value for regression coefficient z-test; ci: confidence interval; Q_M_-Test: test for subgroup differences; p_Q_: p-value for test for subgroup differences; risk of bias: as assessed by the Hulsbosch Ratings; SST validity: stop-signal task validity; overall quality: risk of bias and SST validity ratings combined; Setting: setting of recruitment for ADHD group.

| Moderator | *B* (SE) | *z* | *p* | *ci* | *Q<sub>M</sub>-Test* | *p<sub>Q</sub>* |
| --- | --- | --- | --- | --- | --- | --- |
| Risk of bias |  |  |  |  | *Q<sub>M</sub>*(2) = 0.260 | .878 |
| High (*k* = 13) | 0.531 (0.116) | 4.583 | <.001 | 0.304, 0.759 |  |  |
| Moderate (*k* = 12) | 0.023 (0.126) | 0.186 | 0.852 | -0.224, 0.271 |  |  |
| Low (*k* = 1) | 0.120 (0.236) | 0.508 | 0.611 | -0.342, 0.582 |  |  |
| SST validity |  |  |  |  | *Q<sub>M</sub>*(2) = 0.593 | .743 |
| Low (*k* = 19) | 0.556 (0.062) | 8.919 | <.001 | 0.434, 0.678 |  |  |
| Moderate (*k* = 5) | -0.036 (0.158) | -0.227 | 0.820 | -0.345, 0.273 |  |  |
| High (*k* = 2) | -0.141(0.186) | -0.760 | 0.447 | -0.505 ,0.223 |  |  |
| Overall quality |  |  |  |  | *Q<sub>M</sub>*(2) = 0.173 | .917 |
| Low (*k* =10) | 0.558 (0.140) | 3.974 | <.001 | 0.283, 0.833 |  |  |
| Moderate/Low (*k* = 12) | -0.010 (0.151) | -0.066 | 0.948 | -0.306, 0.286 |  |  |
| Moderate/High (*k* = 4) | -0.065 (0.189) | -0.343 | 0.732 | -0.435, 0.306 |  |  |
| Comorbidities |  |  |  |  |  |  |
| In Patients |  |  |  |  | *Q<sub>M</sub>*(1) = 0.543 | .461 |
| Allowed (*k* = 19) | 0.508 (0.060) | 8.478 | <.001 | 0.391, 0.626 |  |  |
| Not allowed (*k* = 5) | 0.137 (0.185) | 0.737 | 0.461 | -0.227, 0.500 |  |  |
| In Controls |  |  |  |  | *Q<sub>M</sub>*(1) = 1.592 | .207 |
| Allowed (*k* = 7) | 0.446 (0.097) | 4.584 | <.001 | 0.256, 0.637 |  |  |
| Not allowed (*k* = 16) | 0.157 (0.124) | 1.262 | 0.207 | -0.087, 0.400 |  |  |
| Setting |  |  |  |  | *Q<sub>M</sub>*(2) = 5.432 | .066 |
| Mixed (*k* = 2) | 0.399 (0.085) | 4.698 | <.001 | 0.233, 0.565 |  |  |
| Non-clinical (*k* = 9) | 0.099 (0.123) | 0.805 | 0.421 | -0.142, 0.341 |  |  |
| Clinical (*k* = 11) | 0.259 (0.113) | 2.288 | 0.022 | 0.037, 0.480 |  |  |

### Secondary outcome measures of the SST

Fifteen studies reported the percentage of stop commissions (*Supplementary Figure 4*), only 7 studies reported the percentage of choice errors (*Supplementary Figure 5*) and 9 studies reported omission errors (*Supplementary Figure 6*). Finally, eight of the selected studies provided go accuracy (*Supplementary Figure 7*). The analysis of secondary SST outcome measures revealed no significant differences between patients and controls regarding stop commissions (*g* = 0.142, *p* = 0.064) and choice errors (*g* = 0.242, *p* = 0.079). However, ADHD patients made significantly more omission errors (*g* = 0.418, *p* = 0.01) and had a significantly lower go accuracy (*g* = −0.385, *p* < 0.008). A more detailed description of the results is provided in *Supplementary text 3*.

## Discussion

In this systematic review and meta-analysis, we integrated the data of 27 studies examining the stop-signal task in adult ADHD. The analysis revealed inhibitory control deficits, as expressed in prolonged SSRTs, with a moderate effect size *g* = 0.51. These deficits were not significantly moderated by the study quality, sample characteristics, or clinical parameters. In addition, the analyses of secondary outcome measures revealed greater SST omission errors and reduced go accuracy in patients, although only few (*n* <10) studies were available for these measures.

### Behavioral inhibition deficits in stop-signal response times

The main finding of our meta-analysis is that patients with adult ADHD show reliable moderate deficits in the SSRT. The magnitude of the deficits fits with the outcome of an earlier meta-analysis, which included a much smaller number of studies in adult ADHD (Lipszyc and Schachar, 2010). Our meta-analysis of 27 studies establishes the SSRT as a reliable measure for the assessment of inhibitory control deficits in adult ADHD. Extending previous work, we evaluated the SST quality, using the recommendations set out by a recent consensus paper (Verbruggen et al., 2019) and estimated the risk of bias for each study. The large number of observations enabled us to examine whether study quality, considering RoB and the validity of the SST; demographics (age and gender); IQ or clinical parameters (comorbidities and setting) influence SSRT deficits in patients. To this end, we computed meta-regression and subgroup analyses including all studies for which the respective variables were reported. Surprisingly, none of these variables significantly influenced the magnitude of SSRT deficits in patients. This implies that the prolonged SSRT in patients can be observed in experimental settings even when the study quality and other parameters are not optimal. Taken together, the finding that there were no variables which significantly moderate the SSRT deficits suggests that inhibitory control deficits can be consistently observed and hence, may be a phenotype in adult ADHD.

Another important question is how SSRT deficits relate to clinical symptoms in adult ADHD. In a large-scale study, Kamradt et al. (2014) examined correlations between SSRTs with ratings of current inattentive, hyperactive-impulsive symptoms and executive functions in patients. The study revealed significant moderate relationships between SSRTs and all symptom domains (*r* = 0.23 to 0.30). Using a hierarchical linear regression model, which included other neuropsychological paradigms and demographic covariates, the authors found that only the SSRT and the continuous performance test predicted inattention and hyperactivity-impulsivity total symptom scores. In a similar vein, Stavro, Nigg and colleagues (Nigg et al., 2005; Stavro et al., 2007) also found moderate (*r* = 0.29) relationships between SSRT deficits and executive functions, as expressed in inattentive-disorganized and hyperactive-impulsive symptoms. Thus, inhibitory control deficits, albeit frequently reported in empirical studies, are not well reflected in the diagnostic criteria of adult ADHD. Hence, it could be that these deficits are often neglected during the diagnostic process and therefore also not treated, e.g., in the framework of neurocognitive training.

It is important to note that SSRT deficits are not only found in ADHD but also in other psychiatric disorders such as obsessive-compulsive disorder, addiction or schizophrenia (Lipszyc and Schachar, 2010; Smith et al., 2014). Given the large overlap in inhibitory deficits across disorders, the SST provides no diagnostic value in differentiating these disorders.

Hence, we suggest that the SST could be used for the quantification of inhibitory control deficits in adult ADHD, following the exclusion of other psychiatric disorders in which inhibitory control deficits have been reported. In summary, the finding of reliable moderate deficits in the SSRT implies that the SST could become a valuable tool for the neuropsychological assessment of inhibitory control deficits in adult ADHD. To this end, it would be desirable to collect SST data from large participant samples to obtain normative SSRT distributions, considering age, gender and education. An individual’s performance could be then compared against the respective SSRT distribution to determine their level of performance relative to a normative sample.

### Behavioral inhibition deficits in secondary measures of the SST

In addition to the SSRT, we computed meta-analyses for stop commission errors, go discrimination errors, go omission errors and go accuracy. These analyses revealed small to moderately greater omission errors (*g* = 0.418) and reduced go accuracies (*g* = −0.385) in patients. However, only a few studies have reported omission errors (*n* = 9) or go accuracy (*n* = 8), and thus, these findings should be interpreted as preliminary evidence.

For omission errors, the study with the largest reported effect size (*g* = 0.78) was conducted by Roberts et al. (2011). In this study, 30 adult patients with ADHD and 28 control subjects participated in a classical SST paradigm (Logan et al., 1984). Contrary to Roberts et al. (2011), an even negative albeit not significant effect (*g* = −.17) was reported by Bialystok et al. (2017). In their study monolingual and bilingual patients (*n* = 28 monolingual, *n* = 28 bilingual) and controls (*n* = 36 monolingual, *n* = 37 bilingual) participated in a slightly modified version of the SST. Hence, albeit there was some variance in effect sizes across the studies included in the analysis, there were on average significant small to moderate group differences in omission errors.

A similar variability was also observed for accuracy in go trials, where the largest group differences (*g* = −0.64) were reported by Epstein et al. (2001) and the smallest group differences (*g* = 0.24) were observed by Szekely et al. (Szekely et al., 2017; fMRI study). However, this latter study can be considered as an outlier and a meta-analysis excluding this study led to an increased *g* = −0.489. In summary, there is some evidence that, in addition to the SSRT, omission errors and accuracy in go trials during the SST also reflect neurocognitive deficits in adult ADHD. Since the deficits in omission errors and accuracies are constrained to go trials, they suggest an inability to maintain an ongoing response, which indicates attentional difficulties. This is in line with previous reports of sustained and focused attention deficits in adult ADHD (Marchetta et al., 2008). Further studies should analyze and report the secondary measures or the SST, which could be then submitted to an updated meta-analysis including a higher number of observations.

### Limitations

This review has some limitations. Whilst we used an adapted version of the search syntax proposed by Lipszyc and Schachar (2010) to ensure compatibility with previous reviews, it is possible that the search strategy missed relevant studies due to the exclusion of other terms. To ensure that we detected all studies that fit our selection criteria, we thoroughly scanned the reference lists of the preselected empirical articles, previous meta-analyses and systematic reviews. Secondly, the literature search was restricted to peer-reviewed articles written in English or German. This excluded articles that were unpublished or published in a non-commercial form. Therefore, a publication bias cannot be ruled out. Third, meta-regression analyses based on study-level-averages such as mean age of the overall study sample carry the risk of an ecological bias. For example, within studies, age might be correlated with the outcome (e.g., Congdon et al., 2014), but it might not be across studies, or the other way around (Higgins and Thompson, 2002). For this reason, the possibility that demographic or clinical variables might influence the results of the SST on the individual study level cannot be completely ruled out. Lastly, the quality of most studies included in our meta-analysis was not optimal. Hence, we suggest that future studies should follow recently published best practice recommendations on how to design, implement, analyze and report the SST (Verbruggen et al., 2019) and apply the adapted Hombrados and Waddington criteria to ensure that a representative clinical sample is assessed (Hulsbosch et al. 2021).

## Conclusion

This systematic review and meta-analysis revealed reliable moderate inhibitory control deficits, as reflected in the SST, in adult ADHD. Our meta-regression and subgroup analyses further demonstrated no significant contribution of demographic and study quality variables on the observed group differences in SSRTs. This indicates that inhibitory control deficits may be considered a phenotype in adult ADHD. Our review and meta-analysis suggest that the SST could, in conjunction with other neurocognitive tests and clinical questionnaires, become an important tool for the assessment of inhibitory control deficits in adult ADHD.

## Data Availability

All data produced in the present work are contained in the manuscript.

