## Supplementary Materials for "Assessing inhibitory control deficits in adult ADHD: A systematic review and meta-analysis of the stop-signal task"

We have no conflict of interest to disclose.

* *Send correspondence to:* Prof. Dr. Daniel Senkowski

Dept. of Psychiatry and Neurosciences
Charité Campus Mitte (CCM)
Charité-Universitätsmedizin Berlin
Charitéplatz 1
10117 Berlin, Germany.


#### Supplementary text 1

##### Results of calibration session

In order to increase inter-rater reliability, a calibration session was conducted. This way, the two raters could identify possible sources of disagreement and decide on rules on how to rate ambiguous cases. Moreover, the calibration session revealed a different understanding regarding some of the criteria. Therefore, the following additional rules were applied:

**Representative sample.** We didn't consider a prior diagnosis or medical records if the diagnosis was not confirmed within the study. A judgment by a consultant psychiatrist which is not further described was not considered an interview.

**Sample size.** If the sample size of ADHD patients has a different RoB than the sample size of controls, the overall rating for the sample sizes is oriented towards the category with the higher RoB. For example, if the ADHD sample size is $n$ = 20 (therefore rated as having a moderate RoB) and the sample size of controls however is $n$ = 31 (therefore rated as having a low RoB), the overall rating for sample sizes will be moderate RoB.

**Analysis reporting.** We will not consider whether the method of SSRT estimation was reported, as this aspect is covered by the checklist for SST validity.

**Outcome reporting.** Mean and SD for SSRT must be reported. Studies who did not report SSRT, but kindly provided us with the data after we contacted them, were also rated as having a low RoB in this category. It had to be apparent that non-significant results were equally reported.

**Missing data.** If studies do not explicitly report on missing data, however, it is obvious that data from all participants has been used (e.g., if it can be inferred from the degrees of freedom in the analysis), we rated studies as having a low RoB in this domain, even though the original tool requires a clear statement on missing data.

**Supplementary text 2**

**Assessment of SST validity**

This short checklist is based on the consensus guide by Verbruggen et al. (2019) and focuses only on the validity of the assessment and analysis of the SST.

1. Stop trials

Item**:** Did the task include a sufficient number of stop trials and, furthermore, are stop signals only presented on a minority of trials?

- The task includes 50 or more stop trials AND
- Percentage of stop trials is 25% or less (please see also note).

Note: It is also possible to have a higher percentage of stop signals, additional measures to minimize slowing are required (explicitly instruct participants not to wait and include block-based feedback).

Rating:

- Yes (both conditions fulfilled)
- No (at least one condition is not fulfilled)

2. Tracking procedure

Item: Was the stop-signal delay adapted using a tracking procedure (also called staircase)?

- A tracking procedure was implemented with a sufficient step size (usually 50ms steps are used, 16ms steps are too small).

Rating

- Yes (condition fulfilled)
- No (not fulfilled)

3. Estimation Method for SSRT

Item: Was the integration method used to estimate the SSRT?

- The integration method was used to estimate the SSRT.

Note: There are cases that the integration method is only applied when p(respond|signal) is not about 0.50, otherwise the mean or median method is applied. This case also meets the criterion.

Rating:

- Yes (condition fulfilled)
- No (not fulfilled)

4. Check for invalid estimation of SSRT

Item: SSRT should only be estimated, when the assumptions of the horse race model are not violated. There are different methods to check for non-compliant behaviour, e.g., values for p(respond|signal) were lower than 0.25 or higher than 0.75. Did the researchers inspect behaviour for invalid SSRT estimation for each participant for a potential exclusion?

- A rule for inspection for invalid SSRT estimation was applied for each participant.

Note: There are various methods to identify non-compliant participants like reaction time on unsuccessful stop trials should not be numerically longer than reaction time on go trials. This item is not intended for the exclusion of specific trials within the participants (e.g., exclusion of trials of with reactions shorter than 100ms).

Rating:

- Yes (condition fulfilled)
- No (no condition fulfilled)

Overall Rating of SST validity

- High validity: all four items fulfilled
- Moderate validity: three items fulfilled
- Low validity: two or less items fulfilled

**Supplementary text 3**

**Secondary outcome measures of the SST**

Fifteen studies have reported the percentage of stop commissions. Hedges' $g$ of those studies ranged from -0.234 to 0.660, with 73% of estimates indicating that ADHD patients had a higher stop commission percentage *(Supplementary Figure 4*). The two studies that reported stop commissions but did not use a tracking algorithm displayed larger deviations from 50% than the other studies (i.e., Adams et al. 2011: ADHD *M* = 58.2, HC *M* = 62; Marx et al. 2013: ADHD *M* = 42.56, HC *M* = 62). The estimated average Hedges' $g$ based on the random-effects model was $g$ = 0.142 (95% CI: -0.009 to 0.293), which did not significantly differ from zero ($t(14)$ = 2.014, $p$ = 0.064). Moreover, there was no significant heterogeneity ($Q(14)$ = 13.518, $p$ = 0.486, $\hat{\tau}^2$ = 0.002, $I^2$ = 2.754%) with a 95% prediction interval given by -0.040 to 0.324. In addition, there was no indication of outliers as indicated by the studentized residuals (no values larger than ±2.935) and none of the studies could be considered overly influential according to the Cook's distances. Egger's regression test did not indicate funnel plot asymmetry $t(13)$ = 2.037, $p$ = 0.063 *(Supplementary Figure 8).*

Only 7 studies reported the percentage of choice errors, a forest plot is shown in *Supplementary Figure 5.* Hedges' $g$ of those studies ranged from -0.468 to 0.541, with 86% of estimates indicating that ADHD patients made more choice errors. The estimated average based on the random-effects model was $g$ = 0.242 (95% CI: -0.038 to 0.522), which did not significantly differ from zero ($t(6)$ = 2.118, $p$ = 0.079). There was no significant heterogeneity ($Q(6)$ = 7.234, $p$ = 0.299, $\hat{\tau}^2$ = 0.003, $I^2$ = 3.482%). Clark et al. (2007) had a studentized residual larger than ±2.6901 and may be a potential outlier. Cook's distances, however, revealed no overly influential studies. Leaving the study out leads to an average estimate of $g$ = 0.315 (95% CI: 0.125 to 0.506, $t(5)$ = 4.247, $p$ = 0.008), $\hat{\tau}^2$ and $I^2$ decreases to 0. There were not enough studies to test for funnel plot asymmetry, as at least ten studies are recommended for reliable results (Sterne et al., 2011).

A forest plot with 9 studies that reported omission errors is shown in *Supplementary Figure*

*6.* The range of Hedge's $g$ was -0.174 to 0.730, with 78% of estimates hinting that most ADHD patients made more omission errors. The estimated average effect based on the random-effects model was $g$ = 0.418 (95% CI: 0.133 to 0.703), which differed significantly from zero ($t$(8) = 3.379, $p$ = 0.01). The test for heterogeneity reached significance ($Q$(8) = 15.702, $p$ = 0.047, $\hat{\tau}^2$ = 0.077, $I^2$ = 48.319%) and the $I^2$ statistics indicated moderate heterogeneity in the results. For the true outcomes, the 95% prediction interval was -0.282 to 1.118. Bialystok et al. (2017) had a studentized residual larger than ±2.773 and may be a potential outlier as well as potentially over influential according to Cook's distances. Leaving the study out leads to an average estimate of $g$ = 0.524 (95% CI: 0.286 to 0.761, $t$(7) = 5.208, $p$ = 0.001), $\hat{\tau}^2$ decreases to 0.001 and $I^2$ decreases to 1.39. However, there were not enough studies to evaluate funnel plot asymmetry.

Finally, eight of the selected studies provided go accuracy. A forest plot of these studies is shown in *Supplementary Figure 7.* Observed Hedges' $g$ ranged from -0.644 to 0.238, with 88% of estimates indicating that go accuracy was lower for ADHD patients. The estimated average was $g$ = -0.385 (95% CI: -0.635 to -0.136), which significantly differed from zero ($t$(7) = -3.650, p = 0.008). Even though the test for heterogeneity failed to reach significance ($Q$(7) = 9.786, $p$ = 0.201, $\hat{\tau}^2$ = 0.031, $I^2$ = 32.09%), $I^2$ indicated moderate heterogeneity, reflected by a 95% prediction interval between -0.871 and 0.100. The fMRI observation of Szekely et al. (2017) had a studentized residual larger than ±2.734 and may be a potential outlier as well as potentially overinfluential according to Cook's distances. Leaving out this observation increases $g$ to -0.489 (95% CI: -0.608 to -0.368, $t$(6) = -9.963, $p$ < 0.0001) and decreases both $\hat{\tau}^2$ and $I^2$ to 0. Again, funnel plot asymmetry could not be evaluated due to the small number of studies.

#### Supplementary Tables

##### Supplementary Table 1

| | Unadjusted kappa | Adjusted kappa |
|---|---|---|
| Item 1 | 0.920 | 0.923 |
| Item 2 | 1 | 1 |
| Item 3 | 0.752 | 0.769 |
| Item 4 | 0.785 | 0.846 |

*Supplementary Table 1*: Inter-rater reliability for stop signal task validity ratings. Item 1: ≥50 stop trials in total, stop trials constituting ≤25% of all trials; Item 2: staircase algorithm implemented; Item 3: integration method used; Item 4: Cut-Offs applied to ensure valid SSRT estimation.

**Supplementary Table 2**

| | Weighted kappa |
|---|---|
| Equivalent groups | 0.743 |
| Representative sample | 0.792 |
| Sample sizes | 0.861 |
| Selective outcome reporting | 1 |
| Analysis reporting | 1 |
| Missing data | 0.667 |

*Supplementary Table 2:* Inter-rater reliability for RoB ratings. Domains in accordance with the adapted Hombrados and Waddington criteria (Hulsbosch et al., 2021).

**Supplementary Table 3**

| Moderator | | *B (SE)* | *t* | *p* | *ci* | *F*-Test | $p_F$ |
|---|---|---|---|---|---|---|---|
| Age, Sex (*n* = 21) | | | | | | *F*(3,17) = 0.885 | .469 |
| | Intercept | 0.477 (0.054) | 8.872 | <.001 | 0.364, 0.590 | | |
| | Age | 0.123 (0.076) | 1.604 | .127 | -0.039, 0.284 | | |
| | Sex | -0.077 (0.083) | -0.922 | .369 | -0.253, 0.099 | | |
| | Age:Sex | 0.147 (0.126) | 1.161 | .262 | -0.120, 0.413 | | |
| IQ (*n* = 14) | | | | | | *F*(1,12) = 0.123 | .731 |
| | Intercept | 0.564 (0.088) | 6.437 | <.001 | 0.373, 0.755 | | |
| | IQ | 0.006 (0.016) | 0.351 | .731 | -0.029, 0.041 | | |

*Supplementary Table 3:* Meta-regression analyses for SSRT. n: number of studies for which data was available; B: unstandardized regression coefficient. For categorical variables, B is the average estimated effect size for each individual factor level; SE: standard error of regression coefficient; t: t-test for the regression coefficient; p: p-value for regression coefficient t-test; ci: confidence interval; F-Test: test of moderator; $p_F$: p-value for test of moderator; Sex: percentage of males in the individual study samples; IQ: for ADHD and control group combined; Setting: patient setting of ADHD group.

**Supplementary Table 4**

| Moderator | *B (SE)* | *z* | *p* | *ci* | *$Q_M$-Test* | $p_Q$ |
|---|---|---|---|---|---|---|
| Comorbidities | | | | | | |
| In Patients | | | | | $Q_M$ (1) = 0.221 | .639 |
| Allowed (*k* = 18) | 0.517 (0.060) | 8.574 | <.001 | 0.399, 0.635 | | |
| Not allowed (*k* = 3) | -0.083 (0.177) | -0.470 | 0.639 | -0.430, 0.264 | | |
| In Controls | | | | | $Q_M$ (1) = 1.592 | .207 |
| Allowed (*k* = 7) | 0.446 (0.097) | 4.584 | <.001 | 0.256, 0.637 | | |
| Not allowed (*k* = 13) | 0.133 (0.123) | 1.080 | 0.280 | -0.108, 0.373 | | |
| Setting | | | | | $Q_M$ (2) = 4.287 | .117 |
| Mixed (*k* = 2) | 0.399 (0.085) | 4.698 | <.001 | 0.233, 0.565 | | |
| Non-clinical (*k* = 8) | 0.105 (0.136) | 0.771 | 0.441 | -0.162, 0.371 | | |
| Clinical (*k* = 10) | 0.230 (0.112) | 2.057 | 0.040 | 0.011, 0.448 | | |

*Supplementary Table 4*: Subgroup analysis for SSRT. k: number of studies for which data was available; B: regression coefficients (first group is the intercept, for the other groups the coefficients are contrasts); SE: standard error of regression coefficient; Wald-type z-test for the regression coefficient; p: p-value for regression coefficient z-test; ci: confidence interval; $Q_M$-Test: test for subgroup differences; $p_Q$: p-value for test for subgroup differences; risk of bias: as assessed by the Hulsbosch Ratings; SST validity: stop-signal task validity; overall quality: risk of bias and SST validity ratings combined; Setting: setting of recruitment for ADHD group.

#### Supplementary Figures

##### Supplementary Figure 1

| | ADHD | | | Control | | | | | | |
|---|---|---|---|---|---|---|---|---|---|---|
| **Author(s) and Year** | **SSRT Mean** | **SD** | **N** | **SSRT Mean** | **SD** | **N** | **IV, Random, 95% CI** | **Weights** | **g** | **[95% CI]** |
| Aron et al. 2003 | 195 | 55 | 13 | 153 | 27 | 13 | | 7.57% | 0.94 | [ 0.16, 1.72] |
| Cherkasova et al. 2014 | 294.82 | 51.32 | 14 | 244.15 | 34.94 | 12 | | 7.54% | 1.10 | [ 0.32, 1.88] |
| Clark et al. 2007 | 171.6 | 40.3 | 20 | 173.1 | 48.1 | 16 | | 9.29% | -0.03 | [-0.70, 0.63] |
| Crunelle et al. 2013 | 146.2 | 29.4 | 17 | 127.4 | 20.6 | 17 | | 9.04% | 0.72 | [ 0.04, 1.41] |
| Cubillo et al. 2010 | 176 | 157 | 10 | 240 | 196 | 14 | | 7.02% | -0.34 | [-1.17, 0.48] |
| Kamradt et al. 2014 | 260.8 | 73.5 | 170 | 235.8 | 61.5 | 83 | | 19.47% | 0.36 | [ 0.09, 0.62] |
| Marx et al. 2013 | 282.12 | 107.81 | 18 | 239.61 | 66.83 | 20 | | 9.66% | 0.47 | [-0.18, 1.12] |
| Murphy 2002 | 179.36 | 43.23 | 18 | 135.04 | 24.78 | 18 | | 9.36% | 1.23 | [ 0.57, 1.89] |
| Ossmann & Mulligan 2003 | 258 | 101 | 24 | 207 | 43 | 24 | | 11.08% | 0.65 | [ 0.07, 1.22] |
| Pironti et al. 2015 | 187.56 | 76.52 | 20 | 147.02 | 43.02 | 20 | | 9.98% | 0.64 | [ 0.01, 1.27] |
| **RE Model** (Q = 16.52, df = 9, p = 0.06; $I^2$ = 44.2%) | | | | | | | | 100.00% | 0.56 | [ 0.24, 0.88] |

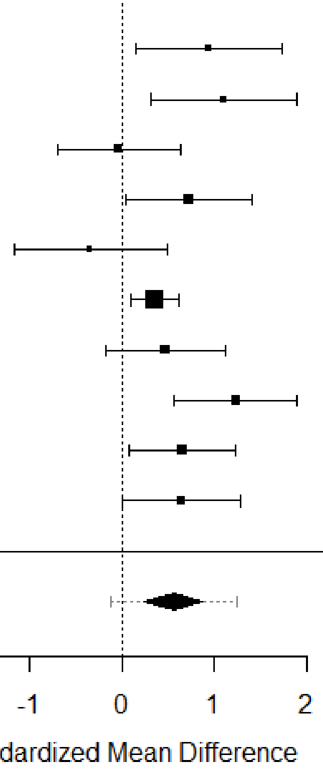

*Supplementary Figure 1:* SSRT for studies with low quality**.** Forest plot showing the observed standardized mean differences (Hedges' g) for SSRT and the estimates of the random-effects model for studies designated as having low quality.

**Supplementary Figure 2**

| | ADHD | | | Control | | | | | |
|---|---|---|---|---|---|---|---|---|---|
| **Author(s) and Year** | **SSRT Mean** | **SD** | **N** | **SSRT Mean** | **SD** | **N** | **Weights** | **g** | **[95% CI]** |
| Adams et al. 2011 | 267.4 | 68.1 | 30 | 229.3 | 50.9 | 27 | 6.99% | 0.62 | [ 0.09, 1.15] |
| Boonstra et al. 2010 | 248.34 | 100.67 | 49 | 191.99 | 67.86 | 49 | 12.05% | 0.65 | [ 0.25, 1.05] |
| Chamberlain et al. 2007 | 235.1 | 73.89 | 20 | 186.5 | 41.14 | 20 | 4.92% | 0.80 | [ 0.17, 1.43] |
| Epstein et al. 2001 | 251.9 | 97.22 | 25 | 209.89 | 42.3 | 30 | 6.71% | 0.57 | [ 0.03, 1.11] |
| Hadas et al. 2021 | 259 | 99 | 52 | 226 | 37 | 49 | 12.41% | 0.43 | [ 0.04, 0.83] |
| Hamzeloo et al. 2018 | 318.09 | 86.11 | 30 | 258.59 | 114.97 | 30 | 7.38% | 0.58 | [ 0.06, 1.09] |
| Lampe et al. 2007 | 239.65 | 115.35 | 16 | 138.18 | 66.86 | 17 | 4.05% | 1.06 | [ 0.36, 1.75] |
| Linhartova et al. 2021 | 294.94 | 102.46 | 26 | 229.08 | 100.03 | 26 | 6.39% | 0.64 | [ 0.09, 1.19] |
| Meachon et al. 2021 | 256.02 | 30.75 | 8 | 254.34 | 43.42 | 19 | 2.78% | 0.04 | [-0.80, 0.88] |
| Nigg et al. 2005 | 251.86 | 67 | 105 | 230 | 52.6 | 90 | 23.84% | 0.36 | [ 0.07, 0.64] |
| Roberts et al. 2011 | 267.41 | 68.1 | 30 | 228.3 | 50.2 | 28 | 7.12% | 0.64 | [ 0.12, 1.17] |
| Sebastian et al. 2012 | 273.89 | 41.2 | 20 | 246.78 | 38.1 | 24 | 5.37% | 0.67 | [ 0.07, 1.28] |
| **RE Model** (Q = 6.84, df = 11, p = 0.81; $I^2$ = 0.0%) | | | | | | | 100.00% | 0.55 | [ 0.42, 0.67] |

**IV, Random, 95% CI**

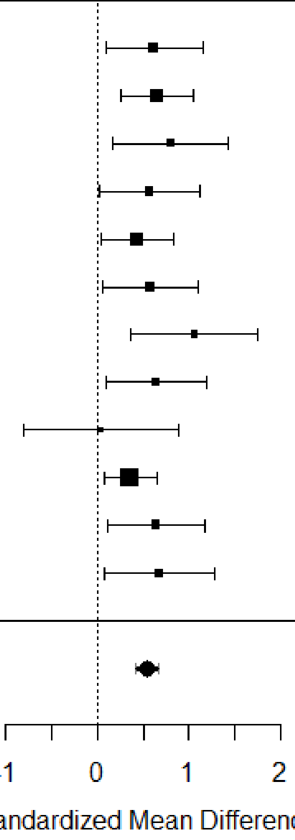

*Supplementary Figure 2:* SSRT for studies with moderate quality**.** Forest plot showing the observed standardized mean differences (Hedges' g) for SSRT and the estimates of the random-effects model for studies designated as having low to moderate quality.

**Supplementary Figure 3**

| | ADHD | | | Control | | | | | | |
|---|---|---|---|---|---|---|---|---|---|---|
| **Author(s) and Year** | **SSRT Mean** | **SD** | **N** | **SSRT Mean** | **SD** | **N** | **IV, Random, 95% CI** | **Weights** | **g** | **[95% CI]** |
| Bekker et al. 2005 | 237.3 | 87.2 | 24 | 185.2 | 38.9 | 24 | | 16.87% | 0.76 | [ 0.18, 1.33] |
| Bialystok et al. 2017 | 333.87 | 78.06 | 50 | 297.39 | 67.84 | 54 | | 35.57% | 0.50 | [ 0.11, 0.89] |
| Congdon et al. 2014 | 195.21 | 70.8 | 25 | 186.38 | 52.64 | 62 | | 24.91% | 0.15 | [-0.32, 0.62] |
| Szekely et al. 2017 MEG | 353.23 | 90.27 | 25 | 296.96 | 79.57 | 46 | | 22.65% | 0.67 | [ 0.17, 1.16] |
| **RE Model** (Q = 3.34, df = 3, p = 0.34; $I^2$ = 3.2%) | | | | | | | | 100.00% | 0.49 | [ 0.09, 0.90] |

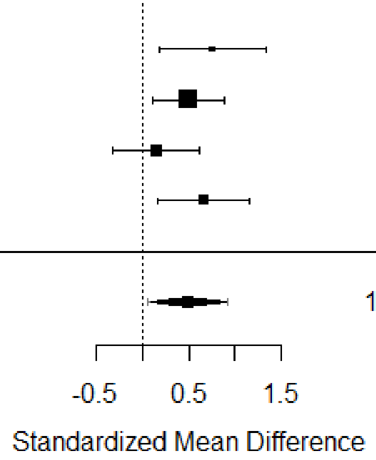

*Supplementary Figure 3:* SSRT for studies with high quality**.** Forest plot showing the observed standardized mean differences (Hedges' g) for SSRT and the estimates of the random-effects model for studies designated as having moderate to high quality.

**Supplementary Figure 4**

| Author(s) and Year | ADHD SC Mean | ADHD SD | ADHD N | Control SC Mean | Control SD | Control N | Weights | g [95% CI] |
|---|---|---|---|---|---|---|---|---|
| Adams et al. 2011 | 58.2 | 22.9 | 30 | 62 | 20.8 | 27 | 7.30% | -0.17 [-0.69, 0.35] |
| Bekker et al. 2005 | 57.24 | 10.58 | 24 | 53.58 | 6.48 | 24 | 6.19% | 0.41 [-0.16, 0.98] |
| Bialystok et al. 2017 | 46.5 | 11 | 50 | 45 | 11 | 54 | 13.01% | 0.14 [-0.25, 0.52] |
| Chamberlain et al. 2007 | 51 | 17 | 20 | 45 | 14 | 20 | 5.18% | 0.38 [-0.24, 1.00] |
| Congdon et al. 2014 | 50.62 | 9.46 | 25 | 50.55 | 5.7 | 62 | 9.09% | 0.01 [-0.45, 0.47] |
| Cubillo et al. 2010 | 47 | 4 | 10 | 46 | 5 | 14 | 3.05% | 0.21 [-0.60, 1.02] |
| Hamzeloo et al. 2018 | 47.67 | 7.87 | 30 | 49.73 | 14.35 | 30 | 7.69% | -0.18 [-0.68, 0.33] |
| Lampe et al. 2007 | 50.06 | 7.13 | 16 | 48.12 | 2.55 | 17 | 4.29% | 0.36 [-0.33, 1.04] |
| Linhartova et al. 2021 | 49.28 | 11.91 | 26 | 42.61 | 7.49 | 26 | 6.70% | 0.66 [ 0.12, 1.20] |
| Marx et al. 2013 | 42.56 | 28.04 | 18 | 32.58 | 38.23 | 20 | 4.92% | 0.29 [-0.35, 0.93] |
| Meachon et al. 2021 | 49 | 2 | 8 | 48 | 3 | 19 | 2.95% | 0.35 [-0.48, 1.18] |
| Ossmann & Mulligan 2003 | 51 | 11 | 24 | 47 | 3 | 24 | 6.19% | 0.49 [-0.08, 1.05] |
| Sebastian et al. 2012 | 48.7 | 6.7 | 20 | 47.58 | 6.3 | 24 | 5.64% | 0.17 [-0.42, 0.76] |
| Szekely et al. 2017 fMRI | 49.06 | 7.37 | 24 | 49.63 | 8.65 | 84 | 9.51% | -0.07 [-0.52, 0.39] |
| Szekely et al. 2017 MEG | 50.98 | 6.44 | 25 | 52.61 | 7.12 | 46 | 8.29% | -0.23 [-0.72, 0.25] |
| **RE Model** (Q = 13.52, df = 14, p = 0.49; $I^2$ = 2.8%) | | | | | | | 100.00% | 0.14 [-0.01, 0.29] |

IV, Random, 95% CI:

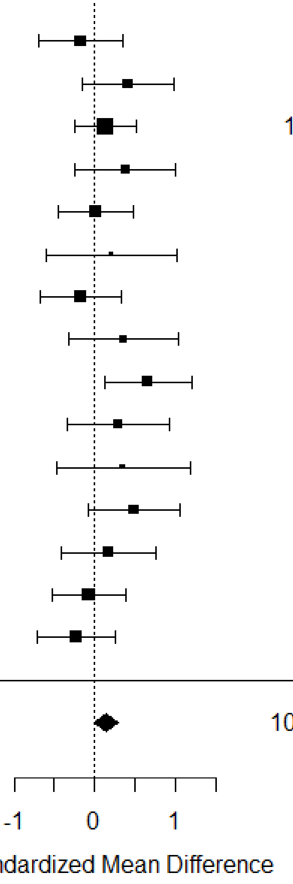

*Supplementary Figure 4:* Forest plot showing the observed standardized mean differences (Hedges' g) for stop commissions (SC, %) and the estimate of the random-effects model.

**Supplementary Figure 5**

| Author(s) and Year | ADHD CE Mean | SD | N | Control CE Mean | SD | N | IV, Random, 95% CI | Weights | g [95% CI] |
|---|---|---|---|---|---|---|---|---|---|
| Adams et al. 2011 | 1 | 1.8 | 30 | 0.52 | 0.8 | 27 | | 15.16% | 0.33 [-0.19, 0.86] |
| Bekker et al. 2005 | 3.43 | 2.7 | 24 | 2.12 | 2.16 | 24 | | 12.88% | 0.53 [-0.04, 1.09] |
| Bialystok et al. 2017 | 4.7 | 5.25 | 50 | 4.15 | 4.85 | 54 | | 26.85% | 0.11 [-0.28, 0.49] |
| Clark et al. 2007 | 3 | 2.8 | 20 | 6 | 8.9 | 16 | | 9.62% | -0.47 [-1.13, 0.19] |
| Linhartova et al. 2021 | 6.43 | 13.68 | 26 | 1.07 | 1.89 | 26 | | 13.91% | 0.54 [-0.00, 1.09] |
| Meachon et al. 2021 | 4 | 4 | 8 | 3 | 2 | 19 | | 6.15% | 0.36 [-0.47, 1.19] |
| Roberts et al. 2011 | 1 | 1.8 | 30 | 0.6 | 0.9 | 28 | | 15.44% | 0.27 [-0.24, 0.79] |
| **RE Model** (Q = 7.23, df = 6, p = 0.30; $I^2$ = 3.5%) | | | | | | | | 100.00% | 0.24 [-0.04, 0.52] |

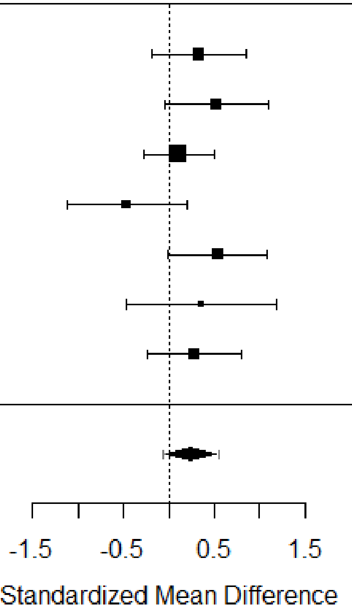

*Supplementary Figure 5:* Forest plot showing the observed standardized mean differences (Hedges' g) for choice errors (CE) in go trials (%) and the estimate of the random-effects model.

**Supplementary Figure 6**

| Author(s) and Year | ADHD OE Mean | SD | N | Control OE Mean | SD | N | IV, Random, 95% CI | Weights | g [95% CI] |
|---|---|---|---|---|---|---|---|---|---|
| Adams et al. 2011 | 3.1 | 4.4 | 30 | 0.74 | 1.1 | 27 | | 12.25% | 0.71 [ 0.18, 1.23] |
| Bekker et al. 2005 | 2.63 | 3.05 | 24 | 1.22 | 1.18 | 24 | | 11.25% | 0.60 [ 0.03, 1.17] |
| Bialystok et al. 2017 | 1.1 | 1.93 | 50 | 1.55 | 3.05 | 54 | | 15.67% | -0.17 [-0.56, 0.21] |
| Lampe et al. 2007 | 4.88 | 5.21 | 16 | 1.88 | 2.98 | 17 | | 9.08% | 0.70 [ 0.01, 1.38] |
| Linhartova et al. 2021 | 7.88 | 10.38 | 26 | 8.51 | 15.67 | 26 | | 11.73% | -0.05 [-0.59, 0.50] |
| Marx et al. 2013 | 13.56 | 17.84 | 18 | 5.58 | 9.14 | 20 | | 9.87% | 0.56 [-0.08, 1.20] |
| Meachon et al. 2021 | 4 | 4 | 8 | 2 | 2 | 19 | | 7.08% | 0.71 [-0.12, 1.55] |
| Roberts et al. 2011 | 3.1 | 4.4 | 30 | 0.7 | 1 | 28 | | 12.37% | 0.73 [ 0.21, 1.25] |
| Sebastian et al. 2012 | 2.53 | 4.3 | 20 | 1.23 | 3.2 | 24 | | 10.69% | 0.34 [-0.26, 0.94] |
| RE Model (Q = 15.70, df = 8, p = 0.05; $I^2$ = 48.3%) | | | | | | | | 100.00% | 0.42 [ 0.13, 0.70] |

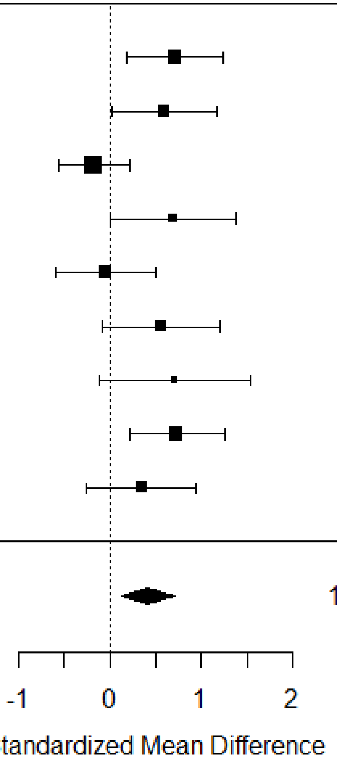

*Supplementary Figure 6:* Forest plot showing the observed standardized mean differences (Hedges' g) for omission errors (OE) in go trials (%) and the estimate of the random-effects model.

**Supplementary Figure 7**

| | ADHD | | | Control | | | | | |
|---|---|---|---|---|---|---|---|---|---|
| **Author(s) and Year** | **ACC Mean** | **N** | **SD** | **ACC Mean** | **SD** | **N** | **Weights** | **g** | **[95% CI]** |
| Congdon et al. 2014 | 97.79 | 6.38 | 25 | 99.18 | 1.76 | 62 | 13.92% | -0.37 | [-0.84, 0.10] |
| Crunelle et al. 2013 | 97.6 | 2.7 | 17 | 98.9 | 1.8 | 17 | 8.13% | -0.55 | [-1.23, 0.12] |
| Epstein et al. 2001 | 96.39 | 3.89 | 25 | 98.28 | 1.67 | 30 | 11.60% | -0.64 | [-1.18, -0.11] |
| Hadas et al. 2021 | 89 | 14 | 52 | 94 | 4 | 49 | 17.16% | -0.48 | [-0.87, -0.08] |
| Linhartova et al. 2021 | 85.7 | 19.9 | 26 | 90.42 | 15.28 | 26 | 11.21% | -0.26 | [-0.81, 0.29] |
| Ossmann & Mulligan 2003 | 96 | 4 | 24 | 98 | 3 | 24 | 10.58% | -0.56 | [-1.13, 0.01] |
| Szekely et al. 2017 fMRI | 91.78 | 8.36 | 24 | 89.43 | 10.17 | 84 | 14.36% | 0.24 | [-0.22, 0.69] |
| Szekely et al. 2017 MEG | 80.67 | 8.56 | 25 | 86.65 | 10.5 | 46 | 13.06% | -0.60 | [-1.09, -0.11] |
| **RE Model** (Q = 9.79, df = 7, p = 0.20; $I^2$ = 32.1%) | | | | | | | 100.00% | -0.39 | [-0.63, -0.14] |

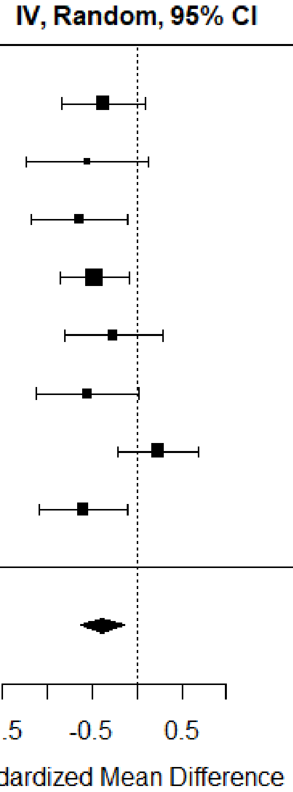

*Supplementary Figure 7:* Forest plot showing the observed standardized mean differences (Hedges' g) for accuracy (ACC) in go trials (%) and the estimate of the random-effects model.

**Supplementary Figure 8**

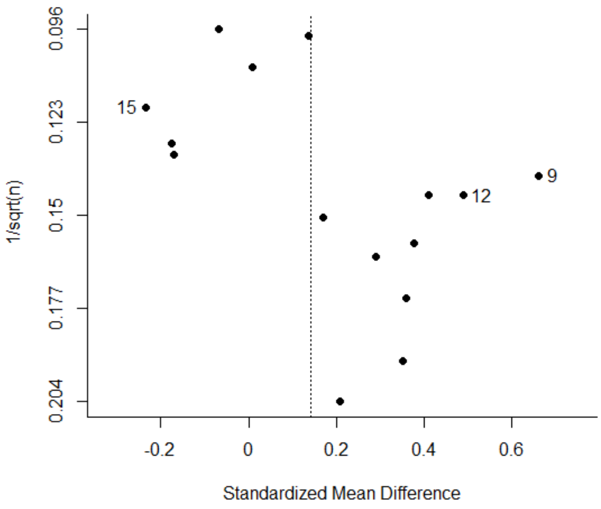

*Supplementary Figure 8:* Funnel plot for stop commissions plotting SMDs against the inverse of the square root of the sample size. [9]Linhartova et al. (2021); [12]Ossman & Mulligan (2003); [15]Szekeley et al. (2017), MEG version.

Crunelle, C.L., Veltman, D.J., van Emmerik-van Oortmerssen, K., Booij, J., van den Brink,

W., 2013. Impulsivity in adult ADHD patients with and without cocaine dependence. Drug Alcohol Depend. 129, 18–24. doi:https://doi.org/10.1016/j.drugalcdep.2012.09.006

Cubillo, A., Halari, R., Ecker, C., Giampietro, V., Taylor, E., Rubia, K., 2010. Reduced activation and inter-regional functional connectivity of fronto-striatal networks in adults with childhood Attention-Deficit Hyperactivity Disorder (ADHD) and persisting symptoms during tasks of motor inhibition and cognitive switching. J. Psychiatr. Res. 44, 629–639. doi:https://doi.org/10.1016/j.jpsychires.2009.11.016

Epstein, J.N., Johnson, D.E., Varia, I.M., Conners, C.K., 2001. Neuropsychological Assessment of Response Inhibition in Adults With ADHD. J. Clin. Exp. Neuropsychol. 23, 362–371. doi:10.1076/jcen.23.3.362.1186

Hadas, I., Hadar, A., Lazarovits, A., Daskalakis, Z.J., Zangen, A., 2021. Right prefrontal activation predicts ADHD and its severity: A TMS-EEG study in young adults. Prog. Neuro-Psychopharmacology Biol. Psychiatry 111, 110340. doi:https://doi.org/10.1016/j.pnpbp.2021.110340

Hamzeloo, M., Mashhadi, A., Fadardi, J.S., Ghahremanzadeh, M., 2018. Adult attention deficit/hyperactivity disorder among the prison inmates: An investigation of the executive function differences and comorbidity effects. Aust. J. Psychol. 70, 235–245. doi:10.1111/ajpy.12201

Hulsbosch, A.-K., De Meyer, H., Beckers, T., Danckaerts, M., Van Liefferinge, D., Tripp, G., Van der Oord, S., 2021. Systematic Review: Attention-Deficit/Hyperactivity Disorder and Instrumental  Learning. J. Am. Acad. Child Adolesc. Psychiatry 60, 1367–1381. doi:10.1016/j.jaac.2021.03.009

Kamradt, J.M., Ullsperger, J.M., Nikolas, M.A., 2014. Executive function assessment and adult attention-deficit/hyperactivity disorder: tasks versus ratings on the Barkley deficits in executive functioning scale. Psychol. Assess. 26, 1095.

Lampe, K., Konrad, K., Kroener, S., Fast, K., Kunert, H.J., Herpertz, S.C., 2007. Neuropsychological and behavioural disinhibition in adult ADHD compared to borderline personality disorder. Psychol. Med. 37, 1717–1729. doi:10.1017/S0033291707000517

Linhartová, P., Látalová, A., Barteček, R., Širůček, J., Theiner, P., Ejova, A., Hlavatá, P., Kóša, B., Jeřábková, B., Bareš, M., Kašpárek, T., 2020. Impulsivity in patients with borderline personality disorder: a comprehensive  profile compared with healthy people and patients with ADHD. Psychol. Med. 50, 1829–1838. doi:10.1017/S0033291719001892

Linhartová, P., Širůček, J., Ejova, A., Barteček, R., Theiner, P., Kašpárek, T., 2021. Dimensions of impulsivity in healthy people, patients with borderline personality disorder, and patients with attention-deficit/hyperactivity disorder. J. Atten. Disord. 25, 584–595.

Martel, M.M., Nigg, J.T., Schimmack, U., 2017. Psychometrically Informed Approach to

Integration of Multiple Informant Ratings in Adult ADHD in a Community-Recruited Sample. Assessment 24, 279–289. doi:10.1177/1073191116646443

Marx, I., Höpcke, C., Berger, C., Wandschneider, R., Herpertz, S.C., 2013. The Impact of Financial Reward Contingencies on Cognitive Function Profiles in Adult ADHD. PLoS One 8, e67002. doi:10.1371/journal.pone.0067002

Meachon, E.J., Meyer, M., Wilmut, K., Zemp, M., Alpers, G.W., 2021. Evoked Potentials Differentiate Developmental Coordination Disorder From Attention-Deficit/Hyperactivity Disorder in a Stop-Signal Task: A Pilot Study. Front. Hum. Neurosci. 15. doi:10.3389/fnhum.2021.629479

Murphy, P., 2002. Inhibitory control in adults with attention-deficit/hyperactivity disorder. J. Atten. Disord. 6, 1–4.

Nigg, J.T., Stavro, G., Ettenhofer, M., Hambrick, D.Z., Miller, T., Henderson, J.M., 2005. Executive functions and ADHD in adults: evidence for selective effects on ADHD symptom domains. J. Abnorm. Psychol. 114, 706.

Ossmann, J.M., Mulligan, N.W., 2003. Inhibition and Attention Deficit Hyperactivity Disorder in Adults. Am. J. Psychol. 116, 35. doi:10.2307/1423334

Pironti, V.A., Lai, M.-C., Müller, U., Dodds, C.M., Suckling, J., Bullmore, E.T., Sahakian, B.J., 2014. Neuroanatomical Abnormalities and Cognitive Impairments Are Shared by Adults with Attention-Deficit/Hyperactivity Disorder and Their Unaffected First-Degree Relatives. Biol. Psychiatry 76, 639–647. doi:10.1016/j.biopsych.2013.09.025

Roberts, W., Fillmore, M.T., Milich, R., 2011. Linking impulsivity and inhibitory control using manual and oculomotor response inhibition tasks. Acta Psychol. (Amst). 138, 419–428. doi:https://doi.org/10.1016/j.actpsy.2011.09.002

Sebastian, A., Gerdes, B., Feige, B., Klöppel, S., Lange, T., Philipsen, A., Tebartz van Elst, L., Lieb, K., Tüscher, O., 2012. Neural correlates of interference inhibition, action withholding and action cancelation in adult ADHD. Psychiatry Res. Neuroimaging 202, 132–141. doi:https://doi.org/10.1016/j.pscychresns.2012.02.010

Stavro, G.M., Ettenhofer, M.L., Nigg, J.T., 2007. Executive functions and adaptive functioning in young adult attention-deficit/hyperactivity disorder. J. Int. Neuropsychol. Soc. 13, 324–334.

Szekely, E., Sudre, G.P., Sharp, W., Leibenluft, E., Shaw, P., 2017. Defining the neural substrate of the adult outcome of childhood ADHD: a multimodal neuroimaging study of response inhibition. Am. J. Psychiatry 174, 867–876.

van Dongen-Boomsma, M., Lansbergen, M.M., Bekker, E.M., Sandra Kooij, J.J., van der Molen, M., Kenemans, J.L., Buitelaar, J.K., 2010. Relation between resting EEG to cognitive performance and clinical symptoms in adults with attention-deficit/hyperactivity disorder. Neurosci. Lett. 469, 102–106. doi:https://doi.org/10.1016/j.neulet.2009.11.053

Verbruggen, F., Aron, A.R., Band, G.P., Beste, C., Bissett, P.G., Brockett, A.T., Brown, J.W., Chamberlain, S.R., Chambers, C.D., Colonius, H., Colzato, L.S., Corneil, B.D., Coxon, J.P., Dupuis, A., Eagle, D.M., Garavan, H., Greenhouse, I., Heathcote, A., Huster, R.J., Jahfari, S., Kenemans, J.L., Leunissen, I., Li, C.-S.R., Logan, G.D., Matzke, D., Morein-Zamir, S., Murthy, A., Paré, M., Poldrack, R.A., Ridderinkhof, K.R., Robbins, T.W., Roesch, M., Rubia, K., Schachar, R.J., Schall, J.D., Stock, A.-K., Swann, N.C., Thakkar, K.N., Van Der Molen, M.W., Vermeylen, L., Vink, M., Wessel, J.R., Whelan, R., Zandbelt, B.B., Boehler, C.N., 2019. A consensus guide to capturing the ability to inhibit actions and impulsive behaviors in the stop-signal task. Elife 8. doi:10.7554/elife.46323

| Section and Topic | Item # | Checklist item | Location where item is reported |
|---|---|---|---|
| | | populations with only subclinical ADHD symptomatology were not considered.<br>2. Control group: Studies must contain at least one healthy control group.<br>3. Experimental task: Response inhibition performance had to be obtained by the SST or the Change Task, which is a modified version of the SST in which individuals shift to a secondary response after they have inhibited an ongoing response (Verbruggen and Logan, 2009). Studies using atypical SST paradigms such as dual tasks or the selective SST were excluded. The same applies to studies in which participants received feedback on stop-signal performance, because feedback and reward influence response inhibition (Lipszyc and Schachar, 2010; Slusarek et al., 2001).<br>4. Outcome measure: Sufficient test statistics for the stop-signal reaction time (SSRT) must be provided to calculate standardized mean differences (Hedge's g).<br>5. Other criteria: Empirical articles written in English or German language and published or accepted for publication in peer-reviewed journals during the time range 2000-2022." | |
| Information sources | 6 | **Specify all databases, registers, websites, organisations, reference lists and other sources searched or consulted to identify studies. Specify the date when each source was last searched or consulted.**<br>Databases: Medline and PsycInfo (accessed from EBSCOhost). Furthermore, reference lists of the identified empirical articles, previous meta-analysis and systematic reviews were scanned to ensure that all relevant articles were captured. | p. 5 |
| Search strategy | 7 | **Present the full search strategies for all databases, registers and websites, including any filters and limits used.**<br>"To identify relevant articles, an electronic search was conducted up to April 14, 2022 in two major publication databases: Medline and PsycInfo (accessed from EBSCOhost). The following syntax, adapted from Lipszyc and Schachter (2010), was used: [(attention deficit hyperactivity disorder OR ADHD) AND Adult* AND (stop task OR stop signal OR response inhibition OR executive function)]. Limiters were set to only show articles published in peer-reviewed journals since the 1st of January 2000, in English or German. Furthermore, reference lists of the identified empirical articles, previous meta-analysis and systematic reviews were scanned to ensure that all relevant articles were captured." | p. 5 |
| Selection process | 8 | **Specify the methods used to decide whether a study met the inclusion criteria of the review, including how many reviewers screened each record and each report retrieved, whether they worked independently, and if applicable, details of automation tools used in the process.**<br>"The study selection process was conducted by two authors (TZ and DS) and included two stages:<br>1. Initial screening of titles and abstracts using the above-described inclusion and exclusion criteria.<br>2. For the resulting set of studies full texts were obtained and checked in detail for eligibility. Screening of eligible articles was performed in Endnote.<br>In case of discrepancies between authors regarding the eligibility of studies, studies were screened by other team members and disagreements during the first or the second screening process were discussed until consensus was reached." | p. 5 |
| Data collection process | 9 | **Specify the methods used to collect data from reports, including how many reviewers collected data from each report, whether they worked independently, any processes for obtaining or confirming data from study investigators, and if applicable, details of automation tools used in the process.**<br>"Data was extracted by two authors independently (TZ and MS). If statistical values for the meta-analysis were not sufficiently reported, the authors of the articles were contacted and asked to provide the missing information." | p. 5 |
| Data items | 10a | **List and define all outcomes for which data were sought. Specify whether all results that were compatible with each outcome domain in each study were sought (e.g. for all measures, time points, analyses), and if not, the methods used to decide which results to collect**.<br>"The following measures of the SST were extracted from the articles, separately for patients and controls: SSRT as primary outcome; stop | p. 5-6 |

### PRISMA 2020 Checklist

| Section and Topic | Item # | Checklist item | Location where item is reported |
|---|---|---|---|
| | | commission errors (responding on a stop trial); go discrimination errors (e.g., responding with the left arrow key, even though a rightwards pointing arrow was presented); go omission errors (not responding on a go trial) and go accuracy (percentage of correct go trials) as secondary outcomes." | |
| | 10b | **List and define all other variables for which data were sought (e.g. participant and intervention characteristics, funding sources). Describe any assumptions made about any missing or unclear information.**<br>"In addition, important variables that could influence behavioral performance in the planned analysis were extracted and tabulated for each study: Age; IQ; percentage of males; ADHD subtype; years of education; comorbidities; medication status; recruitment setting of patients." | p. 6 |
| Study risk of bias assessment | 11 | **Specify the methods used to assess risk of bias in the included studies, including details of the tool(s) used, how many reviewers assessed each study and whether they worked independently, and if applicable, details of automation tools used in the process.**<br>"Both SST validity and RoB were assessed by two independent raters (TZ and MS). In case of discrepancies between authors, consensus was reached from discussions with other team members. […]<br>For the RoB assessment, we applied the adapted Hombrados and Waddington criteria that have been recently used for studies with ADHD patients (Hulsbosch et al., 2021). […] After all studies were rated regarding SST validity and RoB, a variable of overall study quality combining the RoB ratings and SST validity was created. […] This categorization was used for subgroup analysis (see below)." | p. 6-7 |
| Effect measures | 12 | **Specify for each outcome the effect measure(s) (e.g. risk ratio, mean difference) used in the synthesis or presentation of results.**<br>"Hedges' g was calculated for each individual study and each outcome (primary outcome SSRT and secondary outcomes) displaying the effect size of the group difference." | p.7 |
| Synthesis methods | 13a | **Describe the processes used to decide which studies were eligible for each synthesis (e.g. tabulating the study intervention characteristics and comparing against the planned groups for each synthesis (item #5)).**<br>All studies that matched the eligibility criteria were used for synthesis. | p. 5 |
| | 13b | **Describe any methods required to prepare the data for presentation or synthesis, such as handling of missing summary statistics, or data conversions.**<br>"If statistical values for the meta-analysis were not sufficiently reported, the authors of the articles were contacted and asked to provide the missing information." | p. 5 |
| | 13c | **Describe any methods used to tabulate or visually display results of individual studies and syntheses.**<br>"The results will be visualized using forest plots." | p. 8 |
| | 13d | **Describe any methods used to synthesize results and provide a rationale for the choice(s). If meta-analysis was performed, describe the model(s), method(s) to identify the presence and extent of statistical heterogeneity, and software package(s) used.**<br>"The meta-analysis was carried out in R (version 4.0.3; R Core Team, 2020) and the metafor package (version 3.0.2; Viechtbauer, 2010). Hedges' g was calculated for each individual study and each outcome (primary outcome SSRT and secondary outcomes) displaying the effect size of the group difference. Given that various sources could account for differences in findings across studies, e.g., examination of different patient samples or use of different SST paradigms, a random-effects model was fitted to the data. Instead of the usual large-sample approximation, the sampling variance was adjusted by taking the sample-size weighted average of the Hedges' g values into the equation, as this approach has been shown to be less biased (Lin and Aloe, 2021). For computing confidence intervals, the method introduced by Knapp and Hartung (2003) was chosen. To assess for heterogeneity, (1) $\tau^2$ was estimated using the restricted maximum-likelihood estimator (Viechtbauer, 2005), (2) the Q-test for heterogeneity and (3) the $I^2$ statistics (Higgins and Thompson, 2002) are reported. If heterogeneity between studies is present, i.e., $\tau^2>0$, regardless of whether the Q-test reaches significance, a prediction interval for the true outcomes is provided (Riley et al., 2011). The results will be visualized using forest plots. Furthermore, the model is assessed regarding (1) potential outliers, i.e. studies with studentized residuals larger than the $100\times(1-0.05/(2\times k))$th percentile of a standard normal distribution, considering a Bonferroni correction with $\alpha = 0.05$ (two-sided) for k included studies as well as (2) potentially overinfluential studies, i.e. with a Cook's | p. 7-8, p.10 |

| Section and Topic | Item # | Checklist item | Location where item is reported |
|---|---|---|---|
| | | distance larger than the median plus 6 times the interquartile range of the Cook's distances (Viechtbauer and Cheung, 2010). If outliers were detected, leave-one-out diagnostics for sensitivity analysis were conducted."<br><br>"In addition to the SSRT, the SST reveals other outcome measures that are recommended to be reported (Verbruggen et al., 2019). For the current meta-analysis, we examined stop commission errors, go discrimination errors, go omission errors and go accuracy. All studies which reported these parameters were used for the meta-analyses. The procedures were the same as for SSRT, except that no meta-regression or subgroup analyses were conducted, due to the lower number of available studies." | |
| | 13e | **Describe any methods used to explore possible causes of heterogeneity among study results (e.g. subgroup analysis, meta-regression).**<br>"To assess whether the pre-specified extracted demographic and clinical variables as well as study quality influenced the meta-analytic outcome and to explore the cause of potential heterogeneity, a meta-regression analysis was conducted for continuous (age, sex, IQ) and a subgroup analysis for categorical covariates (RoB, SST validity and overall study quality, comorbidities, patient setting and medication status). […] Finally, for both meta-regressions and subgroup analyses an omnibus test of moderators was conducted, testing all coefficients excluding the intercept against 0. If the omnibus test reaches significance, this might be an indication that some of the heterogeneity could be explained by the predictors included in the model (Viechtbauer, 2010)." | p. 9-10 |
| | 13f | **Describe any sensitivity analyses conducted to assess robustness of the synthesized results.**<br>"If outliers were detected, leave-one-out diagnostics for sensitivity analysis were conducted (Viechtbauer, 2010)." | p. 8 |
| Reporting bias assessment | 14 | **Describe any methods used to assess risk of bias due to missing results in a synthesis (arising from reporting biases).**<br>"Evidence of publication bias was assessed using a combination of visual and statistical approaches. First, the funnel plot (Copas and Chi, 2000) of standardized mean difference (SMD) against the inverse square root of the sample size was visually inspected for asymmetries (Zwetsloot et al., 2017). […]. However, determination of publication bias using visual inspection methods (such as funnel plots) are often subjective and prone to judgmental errors (Wang and Bushman, 1998). Therefore, it is recommended to additionally compute a quantile-quantile plot (Q-Q plot) to aid in the assessment of publication bias. Next, Egger's regression test was calculated using the inverse of the square root sample size as a predictor to statistically test for asymmetry of the funnel plot (Zwetsloot et al., 2017). Where evidence of bias exists, the trim-and-fill method was applied to adjust for publication bias (Shi and Lin, 2019)." | p. 8-9 |
| Certainty assessment | 15 | **Describe any methods used to assess certainty (or confidence) in the body of evidence for an outcome.**<br>"[…] After all studies were rated regarding SST validity and RoB, a variable of overall study quality combining the RoB ratings and SST validity was created. Studies with high RoB and low SST validity were rated as having low overall quality, studies with moderate or low RoB AND moderate or high SST validity were rated as having moderate to high overall quality. Studies characterized with the remaining combinations of RoB and SST validity (low RoB and low SST validity; moderate RoB and low SST validity; high RoB and moderate SST validity; high RoB and high SST validity) were designated to the category moderate to low overall quality. This categorization was used for subgroup analysis (see below)." | p. 7 |
| **RESULTS** | | | |
| Study selection | 16a | **Describe the results of the search and selection process, from the number of records identified in the search to the number of studies included in the review, ideally using a flow diagram.**<br>"The electronic search resulted in 1186 articles in MEDLINE and 1353 in PsycInfo (Figure 1). […] The screening of reference lists revealed no further articles. Finally, there were a total of 26 publications with 27 studies included in the meta-analysis (1799 participants; ADHD = 883; controls = 916). " | p. 10-11, p. 35 |
| | 16b | **Cite studies that might appear to meet the inclusion criteria, but which were excluded, and explain why they were excluded.**<br>"In six studies not sufficient statistical values for the meta-analysis were reported and authors were contacted. We received data from four studies, which were then included in the final sample. Finally, it is important to note that Bekker et al. (2005a, 2005b) and van Dongen- | p. 11 |

| Section and Topic | Item # | Checklist item | Location where item is reported |
|---|---|---|---|
| | | Boomsa et al. (2010) reported data from the same experimental session and the same participant sample. The same accounted for Nigg et al. (2005), Stavro et al. (2007) and Martel et al. (2017) as well as for Linhartová et al. (2020) and Linhartová et al. (2021). For these groups of articles, the reported values were extracted and counted as one study in the meta-analysis. Finally, Szekely et al. (2017) conducted two SST experiments, one implemented for fMRI and one for MEG. Even though samples for these two experiments partially overlap (63 completed the SST during MEG and fMRI, 85 during fMRI only, and 33 during MEG only), they were treated as individual observations in the analysis." | |
| Study characteristics | 17 | **Cite each included study and present its characteristics.**<br>"Sample characteristics for all included studies are found in Table 1." | p. 11, p. 26-28 |
| Risk of bias in studies | 18 | **Present assessments of risk of bias for each included study.**<br>"SST validity was evaluated for all 26 articles across 4 items, resulting in 104 individual ratings. Table 3 provides an overview of the ratings. […] RoB was evaluated for all 26 articles across 6 domains, resulting in 156 individual ratings. Table 5 provides an overview of the RoB ratings. […]" | p. 12, p. 30-32 |
| Results of individual studies | 19 | **For all outcomes, present, for each study: (a) summary statistics for each group (where appropriate) and (b) an effect estimate and its precision (e.g. confidence/credible interval), ideally using structured tables or plots.**<br>"Figure 2 presents the forest plot of the observed group differences in the SSRT for 27 observations.<br>Fifteen studies reported the percentage of stop commissions (Supplementary Figure 4), only 7 studies reported the percentage of choice errors (Supplementary Figure 6) and 9 studies reported omission errors (Supplementary Figure 7). Finally, eight of the selected studies provided go accuracy (Supplementary Figure 8)." | p. 36-37<br>in supplementary material: p. 13-16 |
| Results of syntheses | 20a | **For each synthesis, briefly summarise the characteristics and risk of bias among contributing studies.**<br>"Twenty-four out of 27 studies prohibited stimulant medication on the day of testing, two studies did not report this information and one study allowed medication (Linhartová et al., 2021). One study tested the effect of stimulant medication on task performance (Chamberlain et al., 2007) and another study allowed medication during testing (Congdon et al., 2014). To maintain similarity between studies, data of those two articles was extracted for the placebo, i.e., the unmedicated group only. Marx et al. (2013) used an SST paradigm that compared performance with and without reward. For this study only data for the non-reward group were extracted. In some articles, information on the presence of comorbidities or the patient setting was ambiguously reported. For example, Aron et al. (2003) report that healthy controls had "no previous contact with psychiatric services" but it is unclear whether potential comorbidities of healthy controls were screened within the study. Therefore, the coding for those two variables might be biased. Upon request, Meachon et al. (2021) provided non-published information on the age and sex distribution in the two groups. Bialystok et al. (2017) provided the means of age and male percentage for the subset that completed the SST. Demographic variables, information about the IQ and other relevant information about the study population were not available for all studies. A summary is given in Table 2." | p. 11-12, p. 29 |
| | 20b | **Present results of all statistical syntheses conducted. If meta-analysis was done, present for each the summary estimate and its precision (e.g. confidence/credible interval) and measures of statistical heterogeneity. If comparing groups, describe the direction of the effect.**<br>"Figure 2 presents the forest plot of the observed group differences in the SSRT for 27 observations. Across studies, Hedges' g values ranged from -0.341 to 1.230. Results of the random-effects meta-analysis revealed a statistically significant moderate average effect size estimate of 0.509 ($t(26) = 7.829$, $p < 0.0001$, 95% CI: 0.376-0.644). Adults with ADHD showed moderately higher SSRTs compared to healthy controls. The I^2 statistic demonstrated moderate evidence of heterogeneity across studies ($Q(26) = 39.546$, $p = 0.043$, τ ̂^2 = 0.030, I^2 = 31.224%). The heterogeneity reflects in a 95% prediction interval ranging between 0.129 and 0.891.<br>According to the Cook's distances, none of the studies was overly influential. However, the study of Szekeley et al. (2017) implementing the SST for fMRI had a studentized residual larger than ±3.113 and, hence, is an outlier in the context of this model. Leaving out this observation would reduce τ^2 to 0.000, I^2 to 0.004% and increase g to 0.524 (95% CI 0.416 to 0.631). Taken together, the random-effects meta-analysis revealed moderate effect sizes (g = 0.51 to 0.52) with larger SSRTs in patients compared to controls. | p.12-13, p.15, in supplementary material: p. 4-5 |

| Section and Topic | Item # | Checklist item | Location where item is reported |
| --- | --- | --- | --- |
| | | Fifteen studies reported the percentage of stop commissions (Supplementary Figure 4), only 7 studies reported the percentage of choice errors (Supplementary Figure 6) and 9 studies reported omission errors (Supplementary Figure 7). Finally, eight of the selected studies provided go accuracy (Supplementary Figure 8). In summary, the analysis of secondary SST outcome measures revealed no significant differences between patients and controls regarding stop commissions (g = 0.142, p = 0.064) and choice errors (g = 0.242, p = 0.079). However, ADHD patients made significantly more omission errors (g = 0.418, p = 0.01) and had a significantly lower go accuracy (g = -0.385, p < 0.008). A detailed description of the results can be found in the supplementary material." | |
| | 20c | **Present results of all investigations of possible causes of heterogeneity among study results.**<br>"Meta-regression analysis was conducted for continuous covariates (age; sex; IQ) and a subgroup analysis for categorical covariates (RoB, SST validity; overall study quality; comorbidities; patient setting; medication status). In this analysis, the data from the fMRI study by Szekeley et al. (2017) were an outlier and therefore, they were excluded from further analysis. […] In summary, our analysis did not reveal variables that significantly moderated the SSRT deficits in adult ADHD." | p. 13-14, p. 33-34 |
| | 20d | **Present results of all sensitivity analyses conducted to assess the robustness of the synthesized results.**<br>SSRT: "According to the Cook's distances, none of the studies was overly influential. However, the observation of Szekeley et al. (2017) implementing the SST for fMRI had a studentized residual larger than ±3.113 and, hence, is an outlier in the context of this model. Leaving out this observation would reduce $tau^2$ to 0.000, $I^2$ to 0.004% and increase g to 0.524 (95% CI 0.416 to 0.631)."<br>Choice errors: "Clark et al. (2007) had a studentized residual larger than ±2.6901 and may be a potential outlier. Cook's distances, however, revealed no overly influential studies. Leaving the study out leads to an average estimate of g = 0.315 (95% CI: 0.125 to 0.506, t(5) = 4.247, p = 0.008), $tau^2$ and $I^2$ decreases to 0."<br>Omission errors: "Bialystok et al. (2017) had a studentized residual larger than ±2.773 and may be a potential outlier as well as potentially over influential according to Cook's distances. Leaving the study out leads to an average estimate of g = 0.524 (95% CI: 0.286 to 0.761, t(7) = 5.208, p = 0.001), $tau^2$ decreases to 0.001 and $I^2$ decreases to 1.39."<br>Go accuracy: "Szekely et al. (2017) had a studentized residual larger than ±2.734 and may be a potential outlier as well as potentially overinfluential according to Cook's distances. Leaving out this observation increases g to -0.489 (95% CI: -0.608 to -0.368, t(6)= -9.963, p < 0.0001) and decreases both $tau^2$ and $I^2$ to 0." | p. 13, in supplementary material: p. 4-5 |
| Reporting biases | 21 | **Present assessments of risk of bias due to missing results (arising from reporting biases) for each synthesis assessed.**<br>"Figure 3A depicts a funnel plot of the studies' SMDs plotted against the inverse of the square root of the sample sizes. Egger's regression test for funnel plot asymmetry was not significant (t(25) = 1.941, p = 0.064). The funnel plot seems to converge close to the average estimate with increasing sample size. A normal quantile-quantile plot is shown in Figure 3B. Most dots in this plot fall inside the 95%-confidence bands. However, in the middle of the line there is a slight skewing to the left, with several dots outside the bands. This is an indication that the data is not perfectly normally distributed and that there could be a subtle publication bias. For exploratory purposes, a funnel plot adjusted for publication bias using the trim-and-fill method was computed (Figure 3C). The adjusted average effect size estimate is g = 0.439, which is still highly significant (t(31) = 6.304, p < 0.0001, 95% CI: 0.297-0.581). Hence, even if the trim-and-fill method is applied, patients show significantly longer SSRTs compared to controls." | p. 13, p.38 |
| Certainty of evidence | 22 | **Present assessments of certainty (or confidence) in the body of evidence for each outcome assessed.**<br>"The analysis of study quality revealed that both RoB assessment and SST validity ratings did not significantly moderate SSRT. For RoB, the estimated effect was largest for studies with low RoB (g = 0.651) and smallest for studies with high RoB (g = 0.531). However, there was only one study designated as having a low RoB, therefore the result for this category should be interpreted with caution. The group of studies assigned low SST validity showed the largest average effect size (g = 0.556), whereas the group rated as having high SST validity showed the smallest average effect size (g = 0.415). There were only two studies with high SST validity, limiting the reliability of the result for this category. Similar to RoB, the study quality did not significantly moderate SSRT, with an effect size g = 0.49 for studies with moderate to high overall quality ratings. Forest plots with subgroups are shown in Supplementary Figures 1-3. In summary, our analysis did not reveal variables that significantly moderated the SSRT deficits in adult ADHD." | p. 14<br>in supplementary material: p. 10-12 |

### PRISMA 2020 Checklist

| Section and Topic | Item # | Checklist item | Location where item is reported |
|---|---|---|---|
| | 24b | **Indicate where the review protocol can be accessed, or state that a protocol was not prepared.**<br>In the PROSPERO database (PROSPERO ID: CRD42021266709). | p. 4 |
| | 24c | **Describe and explain any amendments to information provided at registration or in the protocol.**<br>Not applicable. | |
| Support | 25 | **Describe sources of financial or non-financial support for the review, and the role of the funders or sponsors in the review.**<br>We have no conflict of interest to disclose regarding (financial) support. | p. 1 |
| Competing interests | 26 | **Declare any competing interests of review authors.**<br>We have no conflict of interest to disclose. | p. 1 |
| Availability of data, code and other materials | 27 | **Report which of the following are publicly available and where they can be found: template data collection forms; data extracted from included studies; data used for all analyses; analytic code; any other materials used in the review.**<br>Stop signal task validity rating scale in supplementary material. | In supplementary material p. 2-3 |

*From:* Page MJ, McKenzie JE, Bossuyt PM, Boutron I, Hoffmann TC, Mulrow CD, et al. The PRISMA 2020 statement: an updated guideline for reporting systematic reviews. BMJ 2021;372:n71. doi: 10.1136/bmj.n71

For more information, visit: http://www.prisma-statement.org/

### ICMJE DISCLOSURE FORM

| | |
|---|---|
| **Date:** | 7/9/2022 |
| **Your Name:** | Theresa Ziegler |
| **Manuscript Title:** | Assessing inhibitory control deficits in adult ADHD: A systematic review and meta-analysis of the stop-signal task |
| **Manuscript Number (if known):** | - |

In the interest of transparency, we ask you to disclose all relationships/activities/interests listed below that are related to the content of your manuscript. "Related" means any relation with for-profit or not-for-profit third parties whose interests may be affected by the content of the manuscript. Disclosure represents a commitment to transparency and does not necessarily indicate a bias. If you are in doubt about whether to list a relationship/activity/interest, it is preferable that you do so.

The author's relationships/activities/interests should be defined broadly. For example, if your manuscript pertains to the epidemiology of hypertension, you should declare all relationships with manufacturers of antihypertensive medication, even if that medication is not mentioned in the manuscript.

In item #1 below, report all support for the work reported in this manuscript without time limit. For all other items, the time frame for disclosure is the past 36 months.

| | | **Name all entities with whom you have this relationship or indicate none (add rows as needed)** | **Specifications/Comments (e.g., if payments were made to you or to your institution)** |
|---|---|---|---|
| **Time frame: Since the initial planning of the work** | | | |
| 1 | All support for the present manuscript (e.g., funding, provision of study materials, medical writing, article processing charges, etc.) **No time limit for this item.** | ☒ None | |
| | | | Click the tab key to add additional rows. |
| **Time frame: past 36 months** | | | |
| **2** | Grants or contracts from any entity (if not indicated in item #1 above). | ☒ **None** | |
| **3** | Royalties or licenses | ☒ **None** | |

| | | Name all entities with whom you have this relationship or indicate none (add rows as needed) | Specifications/Comments (e.g., if payments were made to you or to your institution) |
|---|---|---|---|
| **4** | Consulting fees | ☒ **None** | |
| **5** | Payment or honoraria for lectures, presentations, speakers bureaus, manuscript writing or educational events | ☒ **None** | |
| **6** | Payment for expert testimony | ☒ **None** | |
| **7** | Support for attending meetings and/or travel | ☒ **None** | |
| **8** | Patents planned, issued or pending | ☒ **None** | |
| **9** | Participation on a Data Safety Monitoring Board or Advisory Board | ☒ **None** | |
| **10** | Leadership or fiduciary role in other board, society, committee or advocacy group, paid or unpaid | ☒ **None** | |

| | | Name all entities with whom you have this relationship or indicate none (add rows as needed) | Specifications/Comments (e.g., if payments were made to you or to your institution) |
|---|---|---|---|
| **11** | Stock or stock options | ☒ **None** | |
| **12** | Receipt of equipment, materials, drugs, medical writing, gifts or other services | ☒ **None** | |
| **13** | Other financial or non-financial interests | ☒ **None** | |

**Please place an "X" next to the following statement to indicate your agreement:**

☒ I certify that I have answered every question and have not altered the wording of any of the questions on this form.
